# Transcriptomics profiling of Parkinson’s disease progression subtypes reveals distinctive patterns of gene expression

**DOI:** 10.1101/2023.10.11.23296884

**Authors:** Carlo Fabrizio, Andrea Termine, Carlo Caltagirone

## Abstract

Parkinson’s Disease (PD) exhibits significant individual variability, and recent Artificial Intelligence advancements have identified three distinct progression subtypes, each with known clinical features but unexplored gene expression profiles. This study aimed to identify the transcriptomics characteristics of PD progression subtypes, and assess the utility of gene expression data in subtype prediction at baseline. Differentially expressed genes were subtype-specific, and not typically found in other PD studies. Pathway analysis showed distinct and shared features among subtypes. Two had opposing expression patterns for shared pathways, and the third had a more unique profile with respect to the others. Machine Learning revealed that the progression subtype with the worst prognosis can be predicted at baseline with 0.877 AUROC, yet the contribution of gene expression was marginal for the prediction of the subtypes. This study offers insights into PD subtypes transcriptomics, fostering precision medicine for improved diagnosis and prognosis.

## 1 Introduction

Parkinson’s Disease (PD), the prevailing neurodegenerative movement disorder, is experiencing a faster rise in prevalence than other neurological disorders over the last years [1, 2]. The primary pathological feature is the accumulation of misfolded, aggregated α-synuclein in the substantia nigra and other brain regions, which contributes to movement disorders like bradykinesia in combination with either rest tremor, rigidity, or both [3, 4].

PD is a remarkably variable condition, characterized by a wide heterogeneity at individual level, with variations in clinical features, dominant symptoms, and rate of progression [5]. This variability has prompted a number of studies investigating the existence of PD subtypes. To this extent, PD is a well-suited model for precision medicine which, taking individual variability into account, emphasizes fine-grained diagnostics to enhance treatment effectiveness [6]. One of the challenges in PD research is to assign each affected individual to a specific disease cluster, in order to find phenotypic subgroups that may have a particularly good response to specific treatments [3].

While the majority of research concerning data-driven clustering in PD has centred on disease subtyping using baseline cross-sectional data, mounting evidence suggests that PD has a highly heterogeneous progression [7, 8]. Therefore, any static subtype defined at the baseline may not well account for disease progression patterns. Accordingly, PD subtypes instability is particularly pronounced in the early stages of the disease [9, 10] and advanced PD patients exhibit many clinical similarities despite early-stage heterogeneity [11, 12]. The hypothesis of heterogeneous progression in PD found further support in a 2021 study, where a predictive model found that patients show non-sequential, overlapping disease progression trajectories over eight distinct disease states, finally suggesting that static subtype assignment might be ineffective at capturing the full spectrum of PD progression [8].

Recently, α-synuclein Seed Amplification Assays (SAA) resulted as a promising biomarker for the biochemical diagnosis of PD [13], yet this necessitates a cerebrospinal fluid (CSF) sample to be detected, which might not always be readily available. Conversely, peripheral blood is a more accessible sample type and can be subjected to molecular-level analysis, which could provide further details on biomarkers for a finer-grained diagnosis. The identification of disease subtypes in such a complex disease is pivotal to advance therapeutics [14], and RNA-Seq allows for a broad scope view of the biochemical landscape of a specific phenotype [15].

Research on PD blood transcriptomics is consistently highlighting the association of inflammatory pathways, oxidative stress, and mitochondrial processes with the disease [15, 16], also demonstrating that immune cell subtypes play a role in its transcriptomic changes [17]. Nonethe-less, it was noted that RNA-Seq data is often ignored in Machine Learning studies of PD [18], meaning that the potential of this data source remains to be fully exploited.

Efforts in PD progression subtyping research focus on detecting distinct classes of patients based on unique progression patterns, emphasizing the importance of incorporating time as a dimension. Artificial Intelligence algorithms play a crucial role in managing the complexity of time series data, enhancing result reliability, and enabling hypothesis-free experiments.

A pivotal study for PD subtyping employed clustering analysis at baseline and performed a longitudinal evaluation, but it was based on cross-sectional data analysis, thus overlooking the temporal dimension [5]. The most recent attempt in 2022 introduced an intriguing approach, combining NMF-reduced PD representations with Gaussian Mixture Model clustering; however, it lacked a clear temporal framework, resulting in non-overlapping clusters for patients at the latest time point [19]. Contrastingly, a 2019 study by Zhang et al. harnessed a Long Short Term Memory (LSTM) model to identify three PD progression subtypes [20]. LSTM is an AI architecture specifically designed to handle sequential data, such as time series [21]. The analysis of comprehensive clinical and biological data resulted in the identification of three distinct subtypes: in brief, subtype I (S1) starts with mild motor and non-motor symptoms, and motor impairment increases with a moderate rate over time; subtype II (S2) has moderate motor and non-motor symptoms at baseline, with a slow progression rate; subtype III (S3) has significant motor and non-motor symptoms at baseline, and its impairment progresses rapidly over time, thus accounting for a worse prognosis [20]. An improved iteration of this approach, using an LSTM coupled with a Deep Progression Embedding (DPE) model, was shared as a preprint in 2021, aligning with earlier findings but awaiting peer-review [22]. Other authors developed their own algorithm for the identification of progression subtypes [23], but the heterogeneity in the results dependent on the features selected for analysis, and unavailability of clustered subject IDs, made us prone to focusing on PD progression subtypes identified in [20]. Not only the latter is ongoing research, still needing peer-revision for its latest update [22], but has open access to the full analysis code and tables through GitHub.

### 1.1 Aims

To the best of our knowledge, RNA-Sequencing data has never been taken into account in PD progression subtyping research. Although PD subtypes with distinctive progression phenotypes have been identified, their transcriptomics profiles remain unexplored. The present study has two main objectives: (1) to describe the transcriptomics profile of disease progression subtypes, and (2) to subsequently evaluate the usefulness of gene expression data in predicting disease subtype at baseline. The present paper aims to reveal the biological characteristics of disease progression subtypes. We expect to find partially distinct characteristics of gene expression, which should account for the separate identity of the disease subtypes. The identification of unique transcriptomic traits associated with the subtypes may foster precision medicine in PD, with relevant indications for a finer-grained diagnosis and prognosis. Finally, we make available comprehensive results tables and code scripts, fostering the formulation of hypotheses for further experiments on PD subtypes.

## 2 Results

The data preparation process focused on determining which subjects included in the present study (thus meeting the inclusion criterion of having available RNA data) had been clustered into a disease progression subtypes by [20]. Out of the initial 466 PD subjects with assigned subtypes (S1 = 201; S2 = 107; S3 = 158), a total of 407 PD subjects had RNA-Seq data available (S1 = 199; S2 = 52; S3 = 156), and were included in downstream analyses. Outliers’ detection identified 19 records as outliers, and nine subjects showed sex inconsistencies (Supplementary Figure 1). After their removal, the final dataset comprised 2,057 samples from 598 participants. Finally, 58,780 genes were available for the analysis.

### 2.1 Differentially Expressed Genes (DEGs)

Differential expression analysis was conducted to assess changes in gene expression attributable to the progression of the disease over a span of 4 years, thereby incorporating longitudinal measurements for a time course experiment analysis. In particular, each one of the three subtypes was compared to the HC group.

60 DEGs were found for S1 (41 up, 19 down), 34 for S2 (15 up, 19 down), and 32 for S3 (27 up, 5 down). The most part of these DEGs were distinctive of the subtypes, with just six of these DEGs found as shared between two or more subtypes (Figure 1). A list of DEGs with gene names and descriptions, along with the complete results tables from the differential expression analysis, can be found in Supplementary Table 1.

**Figure 1:**
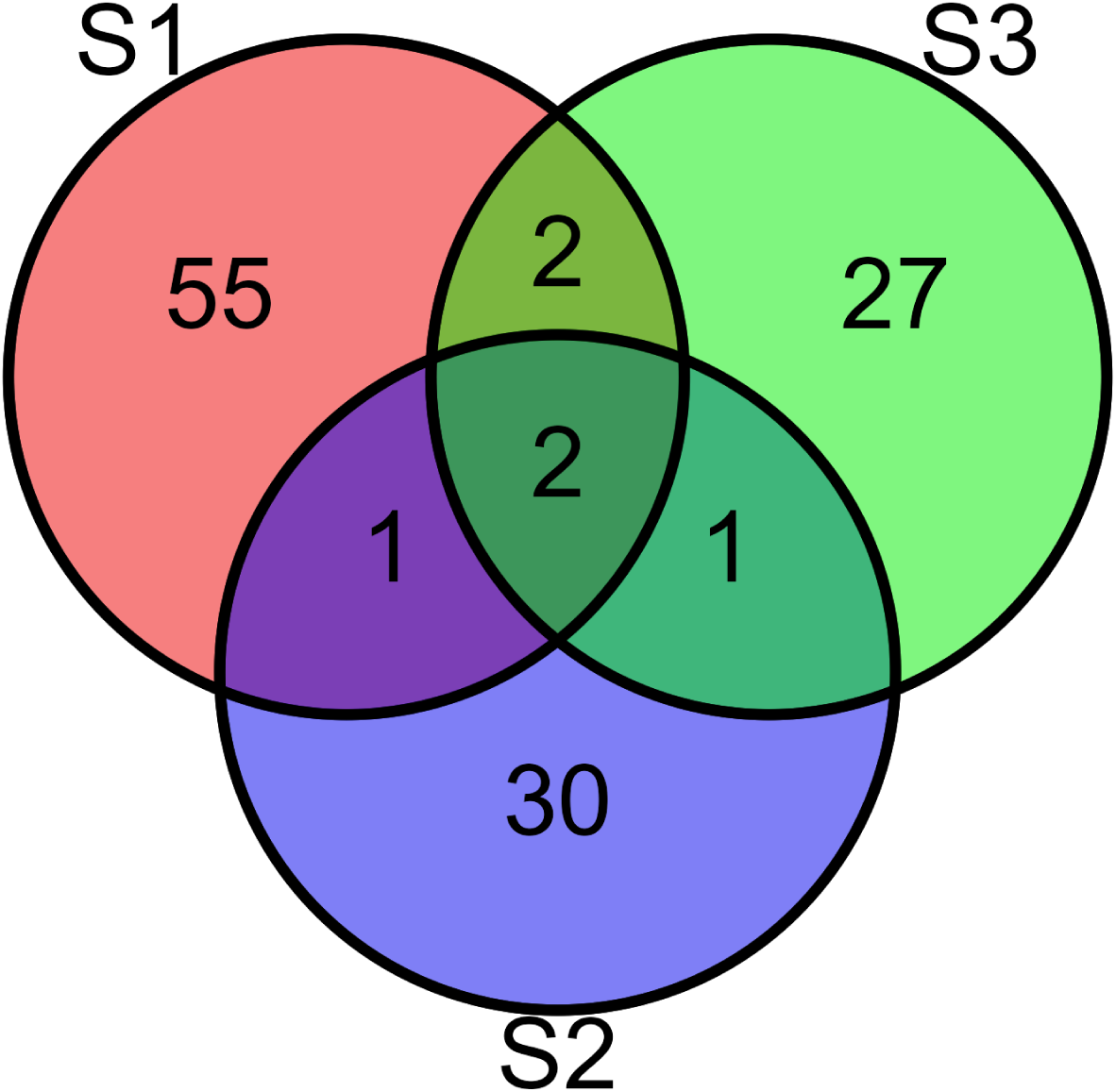
Venn diagram of DEGs for each subtype.

### 2.2 Over Representation Analysis (ORA)

In order to understand the biological pathways associated with the DEGs, ORA was performed on Gene Ontology (GO), Kyoto Encyclopedia of Genes and Genomes (KEGG), and WikiPathways databases. The full list of pathways from the ORA can be found in Supplementary Table 2.

#### 2.2.1 Gene Ontology (GO)

The results include distinctive pathways characterizing each subtype (S1: 73; S2: 18; S3: 16), with seven GO terms in common between S1 and S3, and three in common between S1 and S2 (Figure 2).

**Figure 2:**
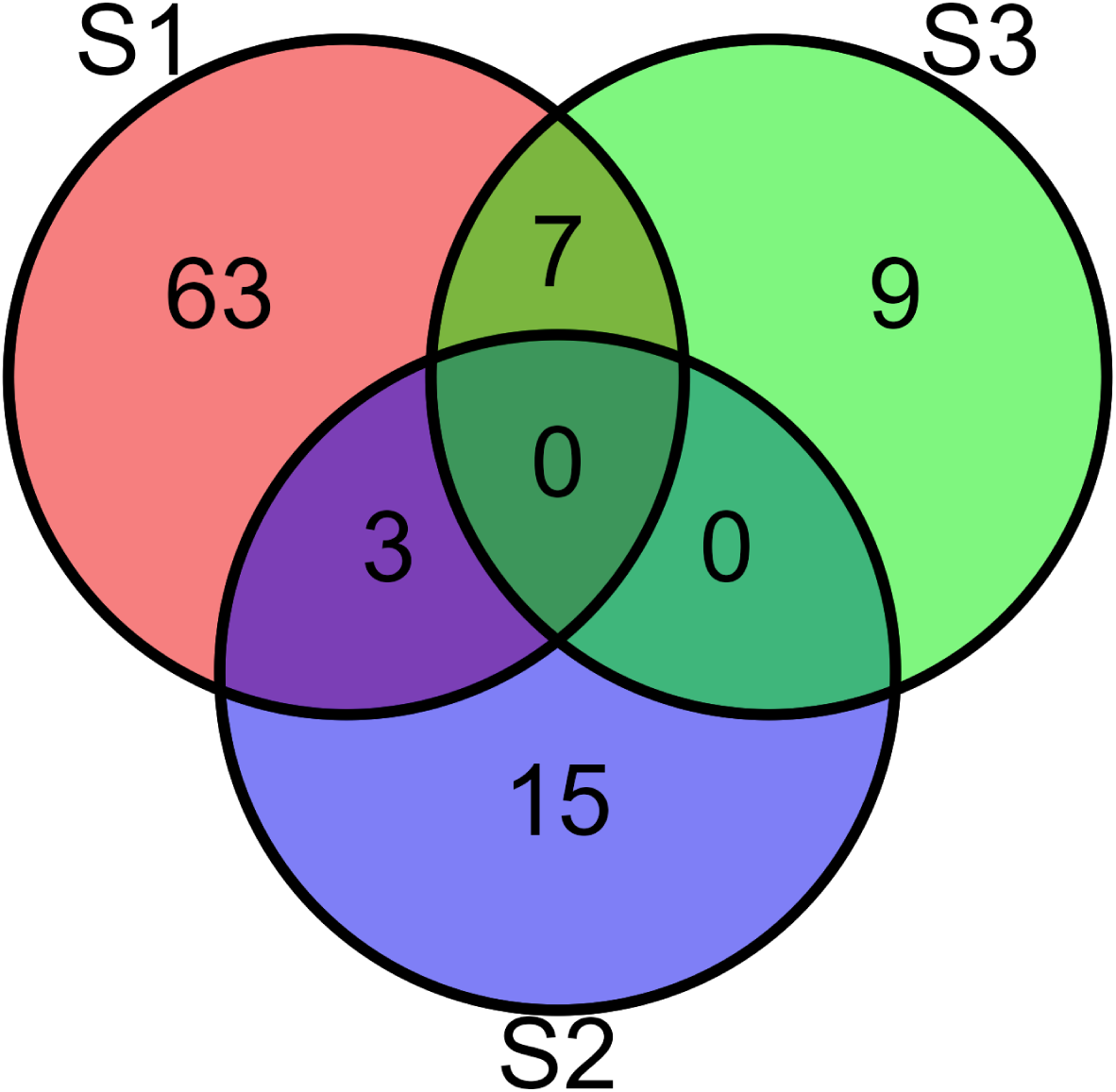
Venn diagram of GO terms for each subtype.

##### 2.2.1.1 S1

The main theme of S1 biological pathways resulting from ORA encompassed cellular energy metabolism, gene expression regulation, and cellular adaptation to various stressors. The presence of pathways associated with *oxidative phosphorylation* (GO:0006119, q-value: 5.54 × 10^-7^), *aerobic respiration* (GO:0009060, q-value: 3.74 × 10^-6^), and *cellular respiration* (GO:0045333, q-value: 1.38 × 10^-5^) indicated a modulation of cellular energy derivation processes, mediated by organic compounds oxidation. Additionally, the presence of pathways related to ATP synthesis, including *mitochondrial ATP synthesis coupled electron transport* (GO:0042775, q-value: 5.54 × 10^-7^), and *proton motive force-driven mitochondrial ATP synthesis* (GO:0042776, q-value: 1.73 × 10^-5^), highlighted the modulation of cellular energy metabolism in this disease sub-type. There were several pathways associated with nucleotide metabolism, including *nucleotide metabolic process* (GO:0009117, q-value: 1.54 × 10^-2^) and *nucleoside phosphate metabolic process* (GO:0006753, q-value: 1.58 × 10^-2^), along with pathways associated with RNA splicing, such as *RNA splicing* (GO:0008380, q-value: 1.14 × 10^-3^), and *mRNA splicing, via spliceosome* (GO:0000398, q-value: 4.13 × 10^-3^). Cellular response to stress pathways were significantly enriched by the set of DEGs, including *cellular oxidant detoxification* (GO:0098869, q-value: 6.68 × 10^-3^), *response to reactive oxygen species* (GO:0000302, q-value: 1.26 × 10^-2^), and *cellular response to toxic substance* (GO:0097237, q-value: 1.26 × 10^-2^).

##### 2.2.1.2 S2

The significantly enriched pathways for this disease subtype mainly pointed to regulation of gene expression and metabolic processes. The most significant term, with the lowest q-value of 7.43 × 10^-7^, was *RNA processing* (GO:0006396). Along with this, several terms associated with RNA metabolism and processing were found significant. These terms included *macromolecule metabolic process* (GO:0043170, q-value: 2.38 × 10^-2^), *RNA metabolic process* (GO:0016070, q-value: 1.23 × 10^-3^), and *nucleic acid metabolic process* (GO:0090304, q-value: 3.51 × 10^-3^). The term with the highest gene ratio found was *metabolic process* (GO:0008152, q-value: 2.92 × 10^-2^), highlighting the modulation of metabolism. This subtype had seven GO terms in common with S1: *RNA splicing, via transesterification reactions; RNA splicing, via transesterification reactions with bulged adenosine as nucleophile; mRNA splicing, via spliceosome*.

##### 2.2.1.3 S3

The main theme of S3 biological pathways resulting from the ORA was the response to oxidative stress and detoxification processes. The pathways with the lower q-values include *response to hydrogen peroxide* (GO:0042542, q-value: 5.76 × 10^-4^), *carbon dioxide transport* (GO:0015670, q-value: 5.76 × 10^-4^), and *oxygen transport* (GO:0015671, q-value: 5.76 × 10^-4^). The presence of these pathways suggests a cellular response to reactive oxygen species, including the catabolic and metabolic processes of hydrogen peroxide. Additionally, pathways related to detoxification processes were highlighted, such as *cellular oxidant detoxification* (GO:0098869, q-value: 6.62 × 10^-3^), *cellular detoxification* (GO:1990748, q-value: 8.13 × 10^-3^), and detoxification (GO:0098754, q-value: 1.08 × 10^-2^). Furthermore, the pathways *cellular response to toxic substances* (GO:0097237, q-value: 8.14 × 10^-3^) and *reactive oxygen species metabolic processes* (GO:0072593, q-value: 2.7 × 10^-2^) further support the main theme of ox-idative stress response and detoxification. This subtype had seven GO terms in common with S1: *response to reactive oxygen species, response to hydrogen peroxide, hydrogen peroxide metabolic process, cellular response to toxic substance, detoxification, cellular oxidant detoxification, cellular detoxification*.

#### 2.2.2 Kyoto Encyclopedia of Genes and Genomes (KEGG)

ORA on the KEGG database showed pathways characterizing each subtype (S1: 16; S2: 1; S3: 2). Most of the pathways resulting from KEGG analysis are unique to the specific subtypes (Figure 3).

**Figure 3:**
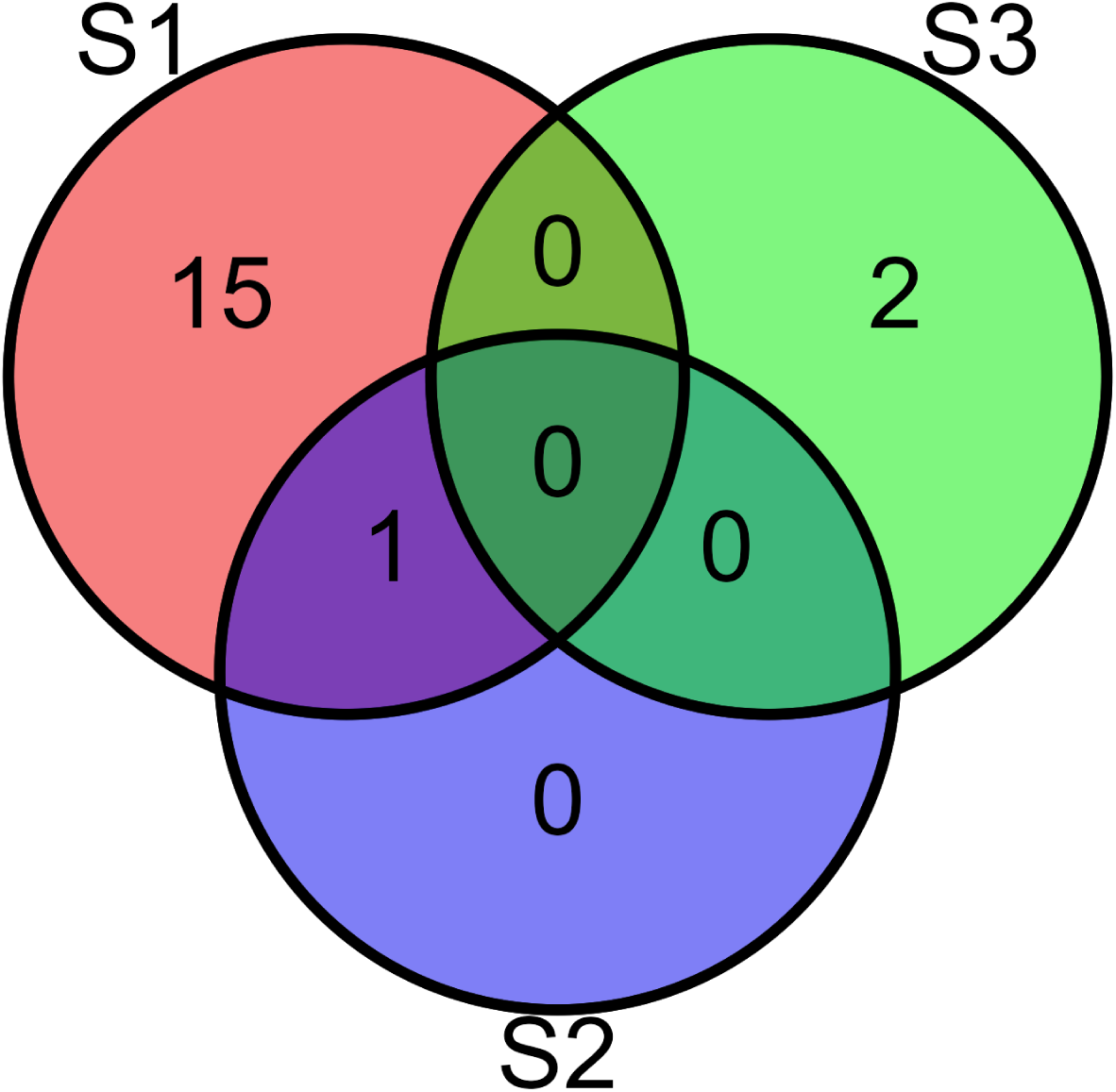
Venn diagram of KEGG terms for each subtype.

##### 2.2.2.1 S1

The main theme of these ORA results on KEGG database is related to neurological diseases and neurodegeneration, including *Parkinson’s disease* (hsa05012, q-value: 1.85 × 10^-5^), *Huntington disease* (hsa05016, q-value: 2.96 × 10^-4^), *prion disease* (hsa05020, q-value: 4.79 × 10^-4^), *amyotrophic lateral sclerosis* (hsa05014, q-value: 2.83 × 10^-3^), and *Alzheimer disease* (hsa05010, q-value: 2.83 × 10^-3^). Additionally, the presence of *oxidative phosphorylation* (hsa00190, q-value: 5.72 × 10^-8^) as the most significant resulting pathway further points to a modulation of energy metabolism.

##### 2.2.2.2 S2

This analysis yielded a single significant pathway, namely *Spliceosome* (hsa03040, q-value: 9.74 × 10^-6^), which further points to the regulation of gene expression. S2 shares his sole pathway with S1 (Figure 3).

##### 2.2.2.3 S3

Here there are two significant pathways, namely *African trypanosomiasis* (hsa05143, q-value: 2.06 × 10^-3^) and *Malaria* (hsa05144, q-value: 2.06 × 10^-3^), suggesting the modulation of pathways involved in detoxification processes.

#### 2.2.3 WikiPathways

ORA on the WikiPathways database showed pathways characterizing each subtype (S1: 5; S2: 2; S3: 19). Most of these pathways are unique to the subtypes (Figure 4).

**Figure 4:**
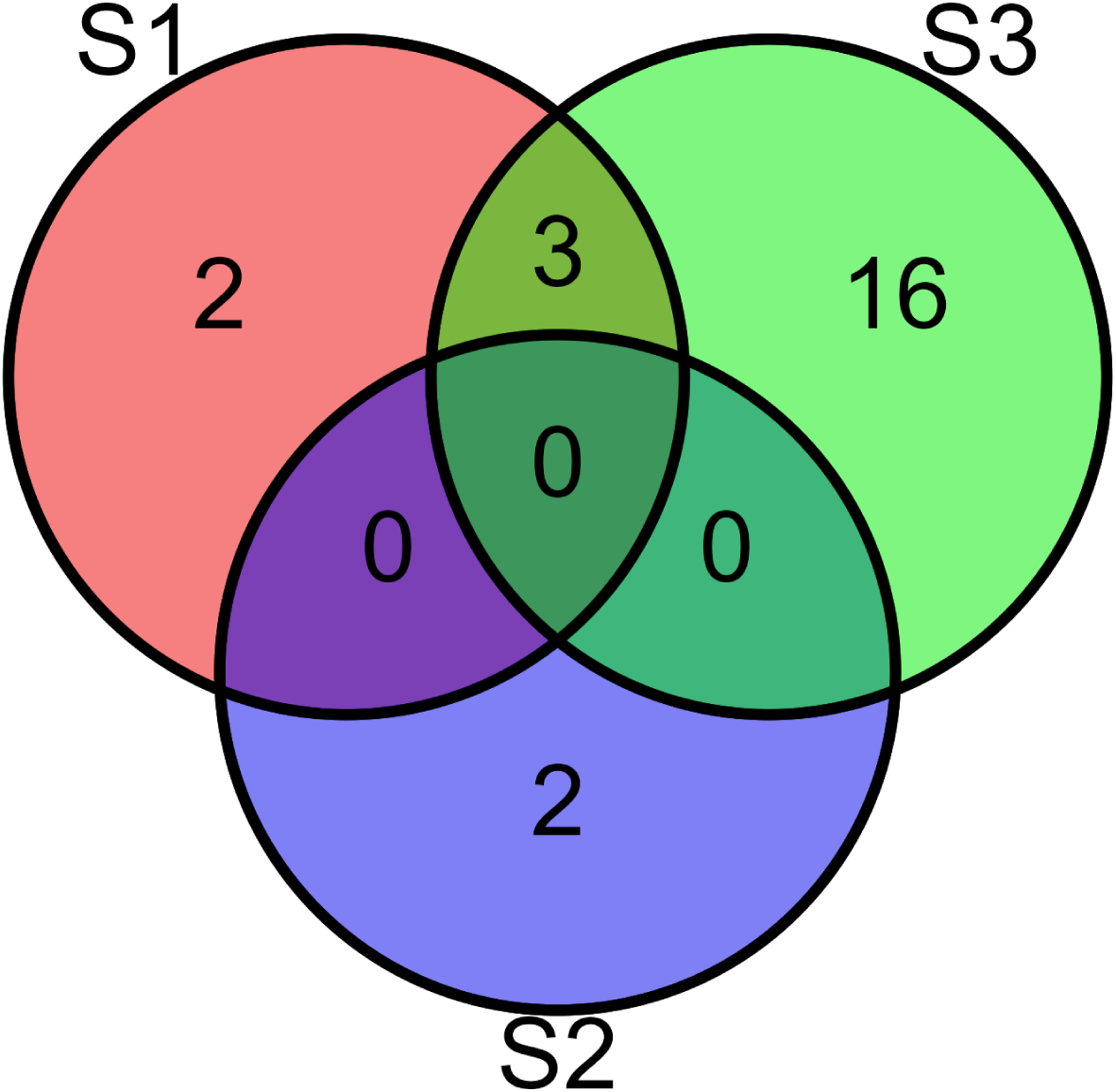
Venn diagram of WikiPathways terms for each subtype.

##### 2.2.3.1 S1

Pathways regarding mitochondrial function and energy production represent the main theme in these results. *Oxidative phosphorylation* (WP623, q-value: 5.68 × 10^-3^) and *Electron transport chain: OXPHOS system in mitochondria* (WP111, q-value: 3.02 × 10^-6^) represent the modulation of ATP generation pathways, along with *Nonalcoholic fatty liver disease* (WP4396, q-value: 1.89

× 10^-3^), associated with mitochondrial dysfunction and impaired energy metabolism.

##### 2.2.3.2 S2

This set of results only includes two pathways, namely *Endoderm differentiation* (WP2853, q-value: 1.85 × 10^-2^) and *Mesodermal commitment pathway* (WP2857, q-value: 1.85 × 10^-2^). Those pathways are enriched by only one gene, namely *NCAPG2*.

##### 2.2.3.3 S3

The main theme in this set of results is related to cellular signaling and metabolism. Many of the terms are pathways involved in signaling processes associated with apoptosis, including *Photodynamic therapy-induced NF-kB survival signaling* (WP3617, q-value: 6.40 × 10^-3^), and *Apoptosis-related network due to altered Notch3 in ovarian cancer* (WP2864, q-value: 6.40 × 10^-3^). Additionally, several pathways are involved in metabolism, including *Vitamin B12 metabolism* (WP1533, q-value: 6.40 × 10^-3^), *Folate metabolism* (WP176, q-value: 6.40 × 10^-3^), and *Selenium micronutrient network* (WP15, q-value: 6.40 × 10^-3^). S3 shares three pathways with S1, namely *Oxidative phosphorylation* (WP623), *Electron transport chain: OXPHOS system in mitochondria* (WP111), and *Mitochondrial complex I assembly model OXPHOS system* (WP4324).

### 2.3 Gene Set Enrichment Analysis (GSEA)

The examination of overall gene expression levels was carried out through GSEA. This analysis is not limited to the set of DEGs, as it accounts for variations in gene expression across all analyzed genes. The complete results tables can be found in Supplementary Table 2.

#### 2.3.1 GO

The number of enriched BP terms found in S1 was 1092, while 1070 enriched terms were found in S2, and 134 enriched terms were found in S3. Venn plots show that most pathways were shared between S1 and S2, while far less were shared with S3 (Figure 5).

**Figure 5:**
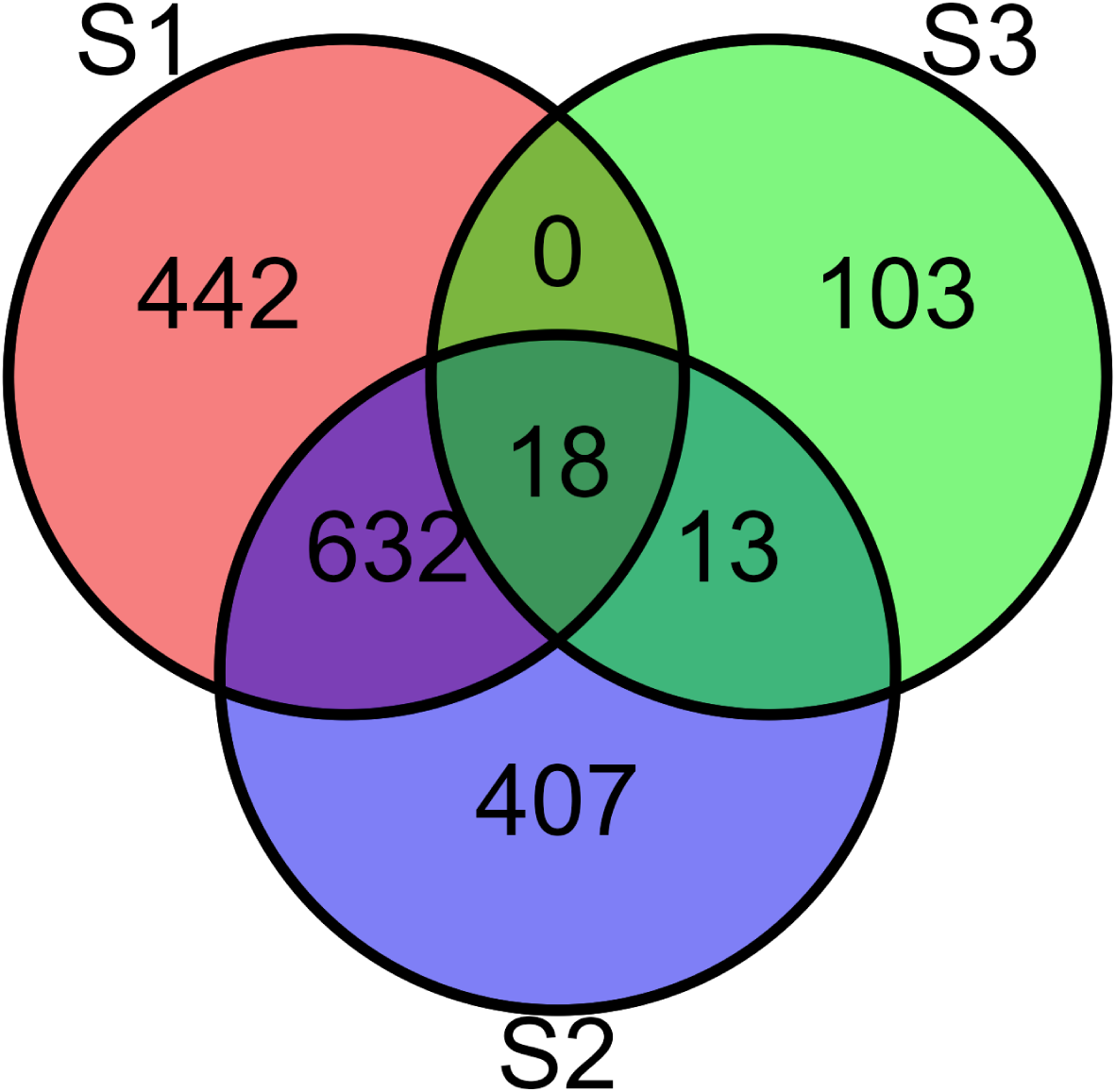
Venn diagram of GO terms for each subtype.

##### 2.3.1.1 S1

Gene Ontology BP domain pathways enrichment revealed main themes related to organismal processes, cell signaling, and energy metabolism. A primary parent term was *Multicellular Organismal Processes* (GO:0032501, q-value: 5.49 × 10^-50^). Also found as enriched with very high significance there were *Nervous System Development* (GO:0007399, q-value: 9.00 × 10^-38^), and *Anatomical Structure Development* (GO:0048856, q-value: 6.30 × 10^-35^). Cell signaling and homeostatic processes were identified as enriched, as exemplified by the presence of *Cell-Cell Signaling* (GO:0007267, q-value: 4.36 × 10^-28^) and *Ion Transmembrane Transport* (GO:0034220, q-value: 5.28 × 10^-21^), along with *Cellular Processes* (GO:0009987, q-value: 7.54 × 10^-12^), and *Cell Communication* (GO:0007154, q-value: 8.86 × 10^-20^). Energy metabolism was mainly represented by *oxidative phosphorylation* (GO:0006119, q-value: 8.56 × 10^-7^), *aerobic respiration* (GO:0009060, q-value: 1.31 × 10^-5^), and *ATP biosynthetic process* (GO:0006754, q-value: 6.13 × 10^-6^)

##### 2.3.1.2 S2

The Gene Ontology analysis revealed biological pathways associated with organismal processes, structures development, and cellular signaling. The most significant pathways were related to *multicellular organismal processes* (GO:0032501, q-value: 5.12 × 10^-68^), followed by development pathways like *nervous system development* (GO:0007399, q-value: 2.26 × 10^-47^) and *anatomical structure morphogenesis* (GO:0009653, q-value: 8.43 × 10^-41^). Signaling pathways included *cell-cell signaling* (GO:0007267, q-value: 1.45 × 10^-29^) and *G protein-coupled receptor signaling pathway* (GO:0007186, q-value: 1.67 × 10^-18^). Pathways related to *response to stimulus* (GO:0050896, q-value: 5.62 × 10^-10^), like *detection of stimulus involved in sensory perception* (GO:0050906, q-value: 9.03 × 10^-13^) were also found in this set of results. Moreover, this GSEA analysis yielded many pathways related to RNA metabolism and processing, such as *positive regulation of transcription by RNA polymerase II* (GO:0045944, q-value: 2.03 × 10^-11^) and *positive regulation of RNA metabolic process* (GO:0051254, q-value: 1.11 × 10^-3^). Pathways for S1 and S2 were mostly shared and related to morphological changes (*nervous system development, anatomical structure development, anatomical structure morphogenesis, tissue development*). Interestingly, all pathways from S1 and S2 showed opposite enrichment scores, indicating that these two groups were characterized by an opposite expression pattern despite sharing most of their enriched pathways (Figure 6).

**Figure 6:**
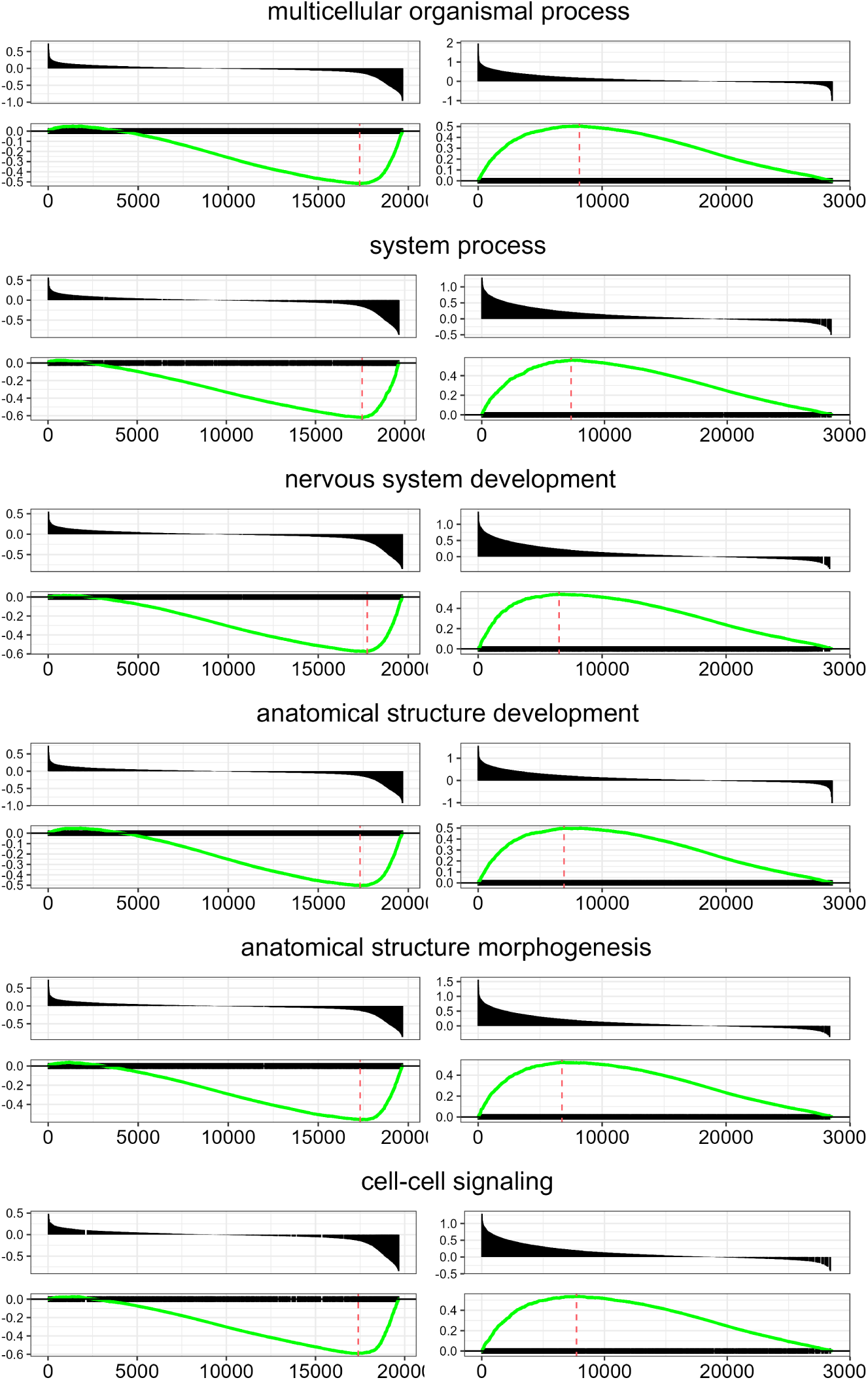
Pathway Enrichment Analysis. Visual representation of six distinct pathways, each labelled with its respective name as a section title. Within each section, there are two sets of plots: S1 on the left and S2 on the right. The upper plots illustrate the positions of gene set members on a rank-ordered list, with the x-axis indicating position and the y-axis representing the ranked list metric. The lower plots display the enrichment scores, with a dashed line indicating the maximum rank of the enrichment score. It is clear to see that all of the represented pathways show opposite enrichment profiles.

##### 2.3.1.3 S3

All enriched pathways in this set of results were distinctive of S3 (none was shared with the other subtypes). One of the prominent themes identified in our analysis is related to sensory perception and signal transduction. Notably, the pathway *detection of chemical stimulus involved in sensory perception of smell* (GO:0050911, q-value: 2.33 × 10^-6^) and *detection of chemical stimulus* (GO:0009593, q-value: 5.96 × 10^-6^) were highly significant in this set of results. Another important theme centers around cell signaling and regulation, with pathways like *positive regulation of antigen receptor-mediated signaling pathway* (GO:0050857, q-value: 3.17 × 10^-5^) and *G protein-coupled receptor signaling pathway* (GO:0007186, q-value: 1.03 × 10^-4^) were highly enriched in this theme, indicating their crucial roles in modulating cellular responses and intercellular communication. Furthermore, our analysis highlighted pathways associated with the regulation of gene expression, such as *regulation of RNA export from nucleus* (GO:0046831, q-value: 9.80 × 10^-5^). Relevantly to Parkinson’s disease, results included *cellular response to misfolded protein* (GO:0071218, 5.92 × 10^-3^) as an enriched pathway.

#### 2.3.2 KEGG

This analysis resulted in 83 enriched terms found for S1, 15 for S2, and 3 for S3 (Figure 7).

**Figure 7:**
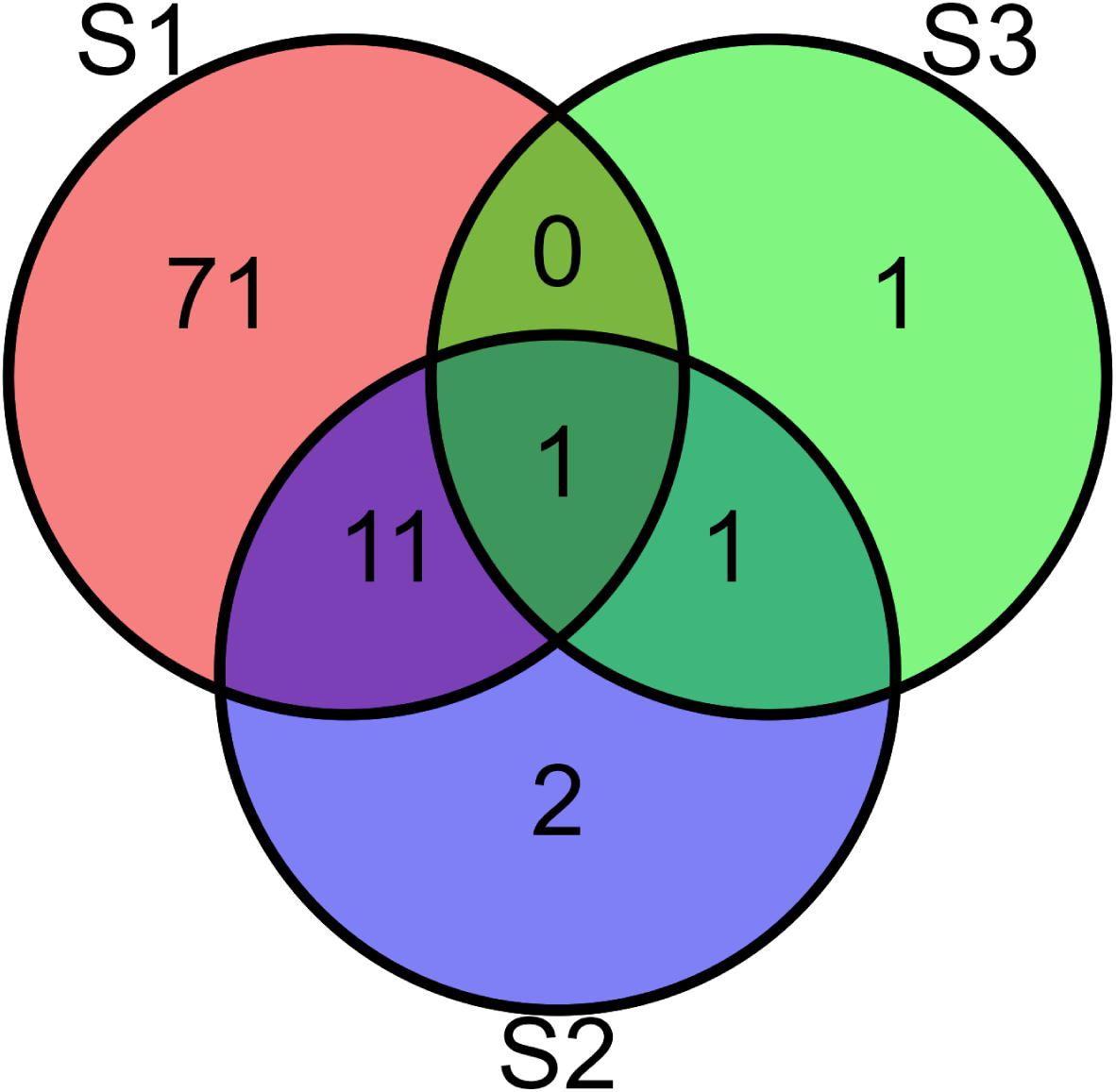
Venn diagram of KEGG terms for each subtype.

##### 2.3.2.1 S1

The main theme of the pathways is the regulation of physiological processes and diseases, including cell signaling and communication pathways, metabolism, disease pathways, along with the regulation of the immune system. The upregulation of protein synthesis is highlighted by the presence of the *Ribosome pathway* (hsa03010, q-value: 2.94 × 10^-14^), and along with *neutrophil extracellular trap formation* (hsa04613, q-value: 1.46 × 10^-13^) and *osteoclast differentiation* (hsa04380, q-value: 1.10 × 10^-6^) pathways, suggest cellular processes and immune system dysregulation. Cell signaling pathways are also significant, such as *glutamatergic synapse* (hsa04724, q-value: 6.77 × 10^-7^), *GABAergic synapse* (hsa04727, q-value: 2.34 × 10^-5^), and *cholinergic synapse* (hsa04725, q-value: 8.77 × 10^-4^) suggesting an implication of disrupted neuronal signaling. Noteworthy, there were again pathways regarding metabolic and energy production dysregulation, such as *oxidative phosphorylation* (hsa00190, q-value: 3.58 × 10^-6^) and *protein digestion and absorption* (hsa04974, q-value: 2.75 × 10^-9^). Finally, pathways related to addiction were found, like *Nicotine addiction* (hsa05033, q-value: 4.38 × 10^-5^) and *Morphine addiction* (hsa05032, q-value: 2.18 × 10^-3^).

##### 2.3.2.2 S2

Pathways involved in cell communication and signal transduction in the nervous system are found modulated in this set of results, including *Neuroactive ligand-receptor interaction* (hsa04080, q-value: 5.93 × 10^-14^), *Calcium signaling pathway* (hsa04020, q-value: 1.85 × 10^-6^), and *Olfactory transduction* (hsa04740, q-value: 2.09 × 10^-8^). Cell development and connectivity was also modulated, as indicated by the presence of pathways like *Wnt signaling pathway* (hsa04310, q-value: 3.98 × 10^-3^) and *Axon guidance* (hsa04360, q-value: 6.84 × 10^-3^). Like in S1, pathways regarding addiction processes were found, such as *Nicotine addiction* (hsa05033, q-value: 1.84 × 10^-3^) and *Morphine addiction* (hsa05032, q-value: 6.16 × 10^-3^).

##### 2.3.2.3 S3

Here three pathways are found as significantly enriched, indicating a positive regulation of *olfactory transduction* (hsa04740, q-value: 6.02 × 10^-10^) along with *Neuroactive ligand-receptor interaction* (hsa 04080, q-value: 2.78 × 10^-10^). Additionally, *Protein export pathway* (hsa03060, q-value: 4.57 × 10^-2^) was found enriched.

#### 2.3.3 WikiPathways

Here 86 enriched terms were found in S1, 40 enriched terms were found in S2, 1 enriched term was found in S3 (Figure 8).

**Figure 8:**
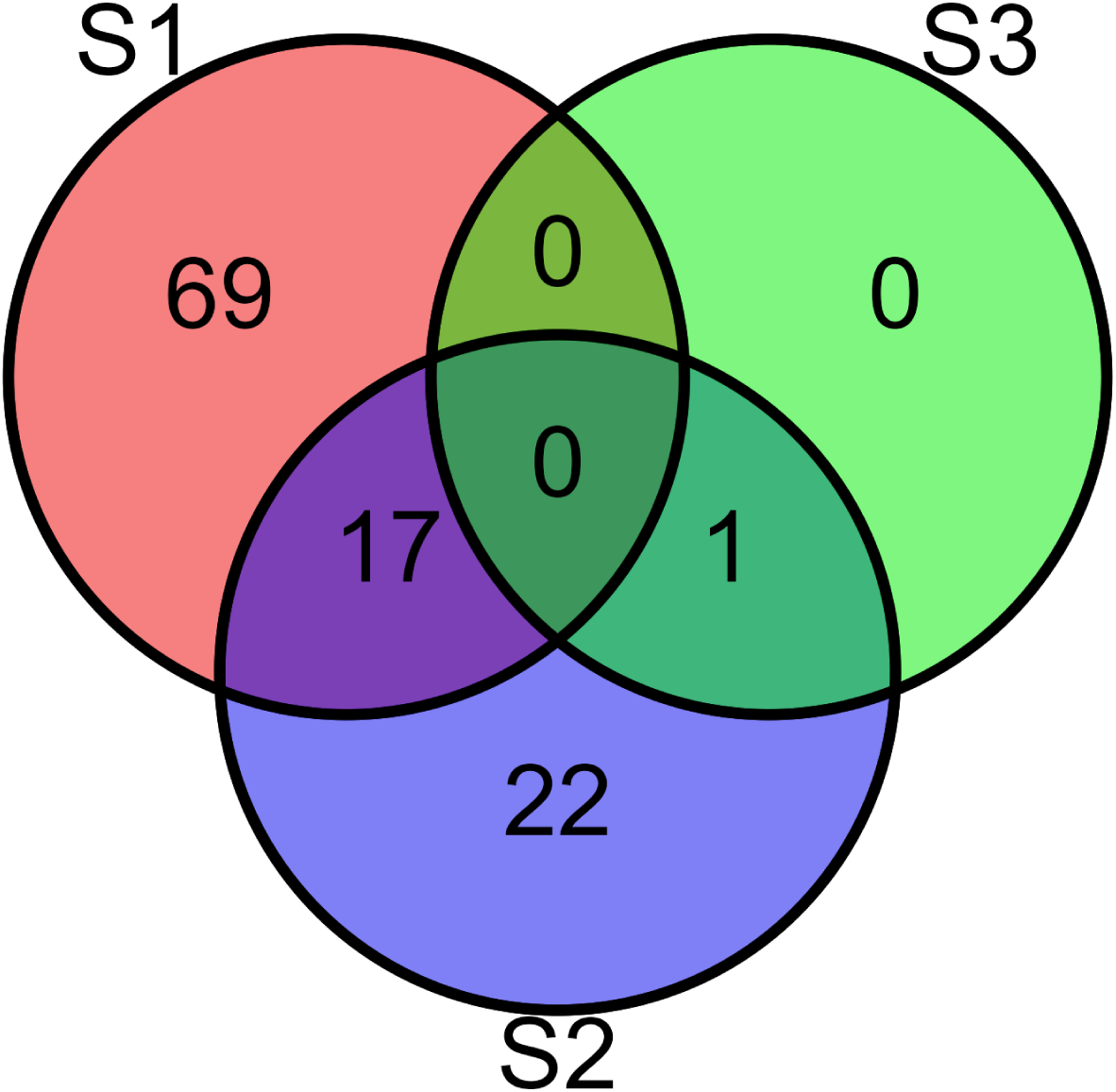
Venn diagram of WikiPathways terms for each subtype.

##### 2.3.3.1 S1

In this set of results, the main theme was driven by enriched pathways related to protein synthesis, cellular metabolism, neuronal signaling, and immune system response. Notably, the *Cytoplasmic ribosomal proteins pathway* (WP477, q-value: 1.81 × 10^-14^) confirmed the modulation of processes related to protein synthesis. Accordingly, the *Electron transport chain: OXPHOS system in mitochondria pathway* (WP111, q-value: 3.58 × 10^−7^) confirmed the importance of oxidative phosphorylation in energy production. The results also included pathways associated with neuronal signaling, like *Phosphodiesterases in neuronal function* (WP4222, q-value: 1.50 × 10^−4^), *mBDNF and proBDNF regulation of GABA neurotransmission* (WP4829, q-value: 1.03 × 10^−2^), and *Neuroinflammation and glutamatergic signaling* (WP5083, q-value: 2.07 × 10^−2^), also pointing out to an involvement of the immune response, along with *IL-3 signaling pathway* (WP286, q-value: 2.65 × 10^−4^). Related to this, the *TYROBP causal network in microglia pathway* (WP3945, q-value: 1.53 × 10^−5^) highlighted the involvement of the regulatory mechanisms within microglia. Another notable theme revolved around disease processes, as there was a significant enrichment of pathways like *Nonalcoholic fatty liver disease* (WP4396, q-value: 1.05 × 10^−3^) and *Hepatitis B infection* (WP4666, q-value: 1.69 × 10^−3^).

##### 2.3.3.2 S2

This set of GSEA results revealed pathways encompassing signaling mechanisms, neurogenesis, developmental processes, immune response, and disease processes. *GPCRs, class A rhodopsin-like* (WP455, q-value: 4.52 × 10^-6^) was the most significant, pointing out to signaling along with *GPCRs, other* (WP117, q-value: 4.70 × 10^-3^), and *GABA receptor signaling* (WP4159, q-value: 4.02 × 10^-2^). Pathways related to cellular differentiation and neurogenesis were also present, such as *dopaminergic neurogenesis* (WP2855, q-value: 1.56 × 10^-2^) and *Neural crest differentiation* (WP2064, q-value: 9.45 × 10^-4^). Developmental processes were represented by pathways such as *cardiac progenitor differentiation* (WP2406, q-value: 2.40 × 10^-4^), *osteoblast differentiation and related diseases* (WP4787, q-value: 2.60 × 10^-4^), and *neural crest differentiation* (WP2064, q-value: 9.45 × 10^-4^). Additionally, results included pathways associated with immune response, such as *host-pathogen interaction of human coronaviruses - interferon induction* (WP4880, q-value: 2.52 × 10^-4^), *immune response to tuberculosis* (WP4197, q-value: 4.34 × 10^-4^), and *SARS coronavirus and innate immunity* (WP4912, q-value: 4.26 × 10^-2^). Finally, we identified pathways associated with genetic disorders and syndromes, including *Prader-Willi and Angelman syndrome* (WP3998, q-value: 1.09 × 10^-2^), *MECP2 and associated Rett syndrome* (WP3584, q-value: 1.35 × 10^-2^), and *Cornelia de Lange Syndrome - SMC1/SMC3 role in DNA damage* (WP5118, q-value: 1.86 × 10^-2^).

##### 2.3.3.3 S3

The GSEA here yielded only one significant pathway, namely *Interactome of polycomb repressive complex 2 (PRC2)* (WP2916, q-value: 4.04 × 10^-2^), indicating a modulation in gene expression regulation and chromatin organization.

### 2.4 Baseline prediction of disease progression subtype

A Machine Learning hierarchical classification approach was implemented to develop a prediction system aimed at identifying the disease subtypes of a newly-diagnosed PD patient, namely at the baseline. Data from multiple modalities were used, including demographics, motor, nonmotor, biospecimen, imaging (See section 4.6, Table 2). The first model in the hierarchy aimed to predict whether the subject was from S3, which has the most distinctive phenotype and is also the most severe. The model achieved a fair performance with 0.814 sensitivity, and 0.757 specificity, yielding an F-Score of 0.828 and a total AUROC of 0.877 (Figure 9).

**Figure 9:**
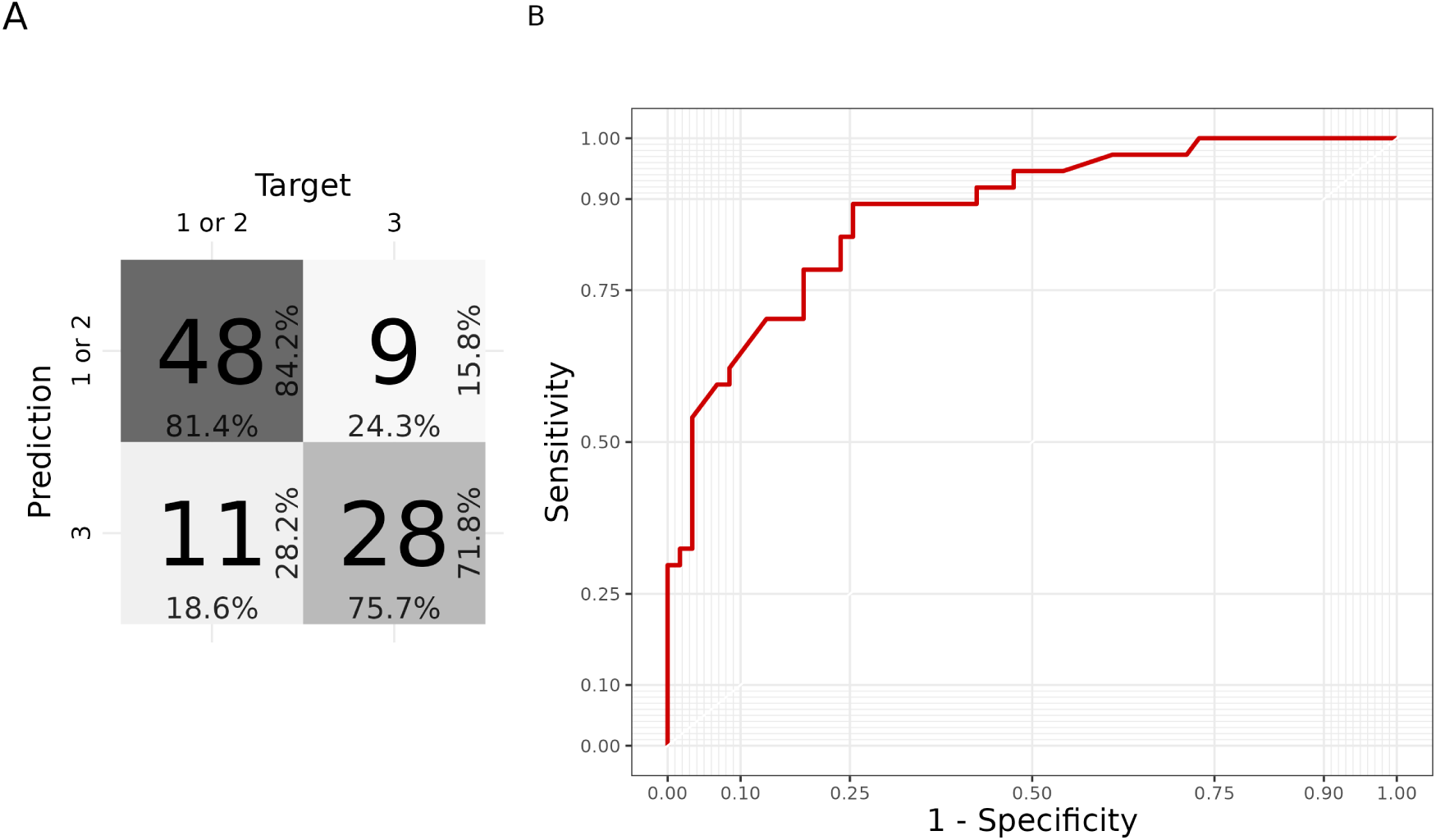
ROC curve and confusion matrix from the first model of the hierarchy.

Variable importance was investigated with the application of an explainable Artificial Intelligence (XAI) method, namely SHAP values. These highlighted the score to MDS-UPDRS Part II (disability evaluation) as the most important factor contributing to S3 identification. Among the most important variables there are other clinical measures, along with a neuroimaging measure (DaTScan Caudate R). Gene expression only had a marginal importance, with low absolute SHAP values, giving little contribution to the final prediction (**??**).

**Figure 10:**
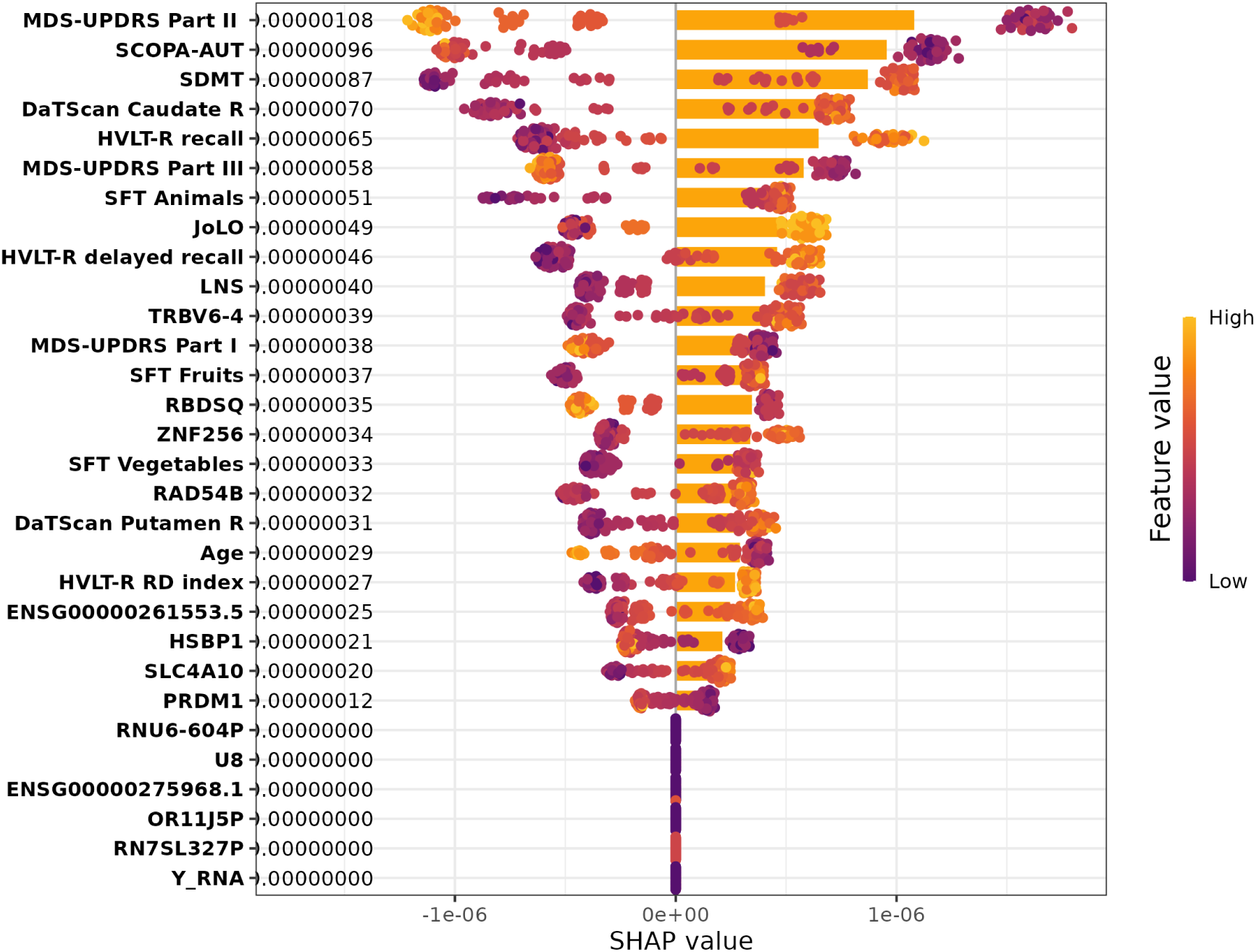
SHAP summary plot representing the contribution of each variable to the prediction of the model.

**??**

For all those subjects that the model did not classify as S3, the second level of the hierarchy included a model aiming to predict whether the subject was from S1 or S2. It achieved a poor performance, with 0.745 sensitivity, 0.25 specificity, yielding a F-Score of 0.77 and a total AU-ROC of 0.576 (Figure 11).

**Figure 11:**
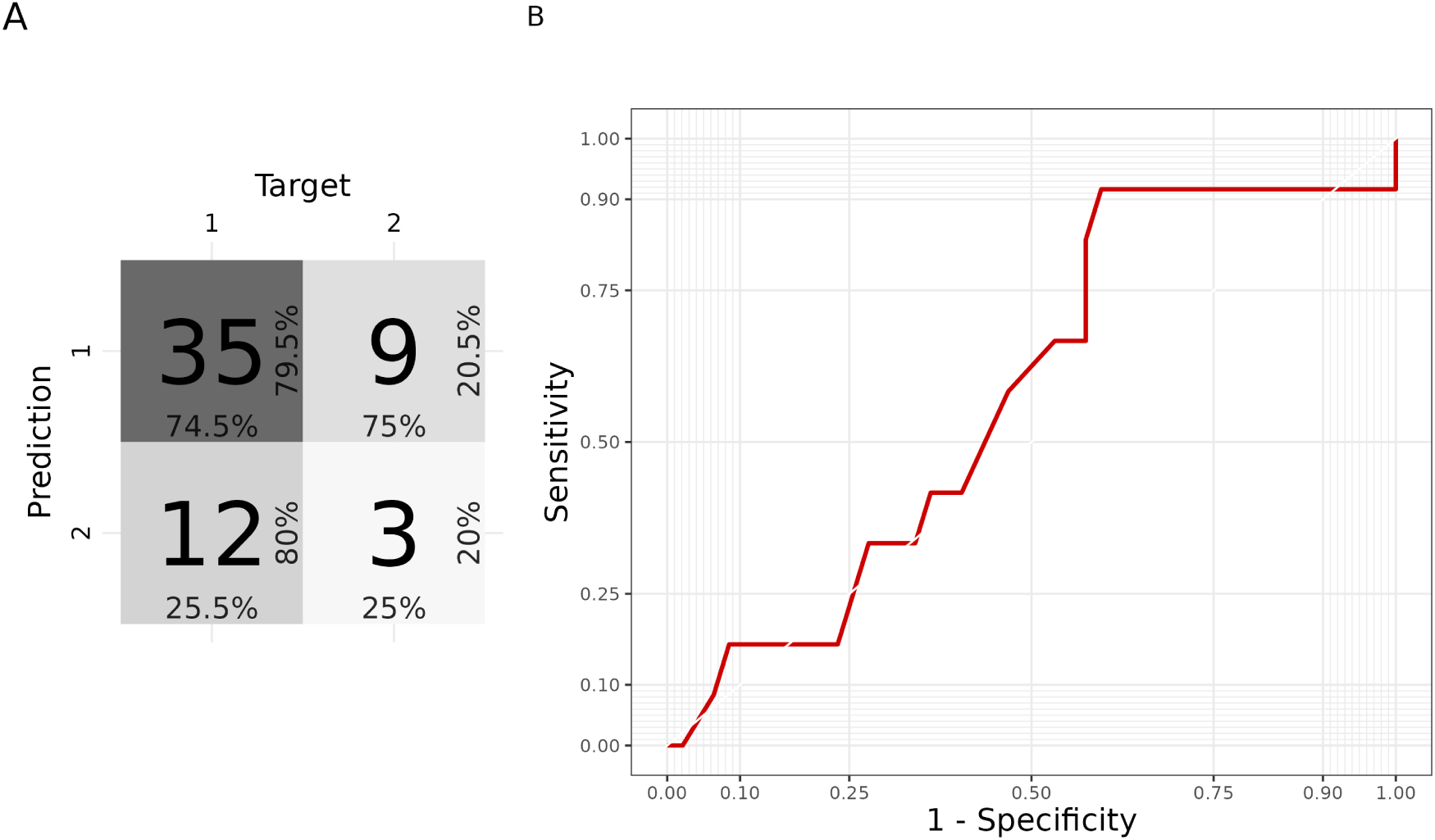
ROC curve and confusion matrix from the second model of the hierarchy.

SHAP values indicated that expression values of *U8, HSBP1, TRBV6-4, and SCL4A10*, along with Benton Judgement of Line Orientation test score, were the most important factors to discriminate between S1 and S2 (Figure 12).

**Figure 12:**
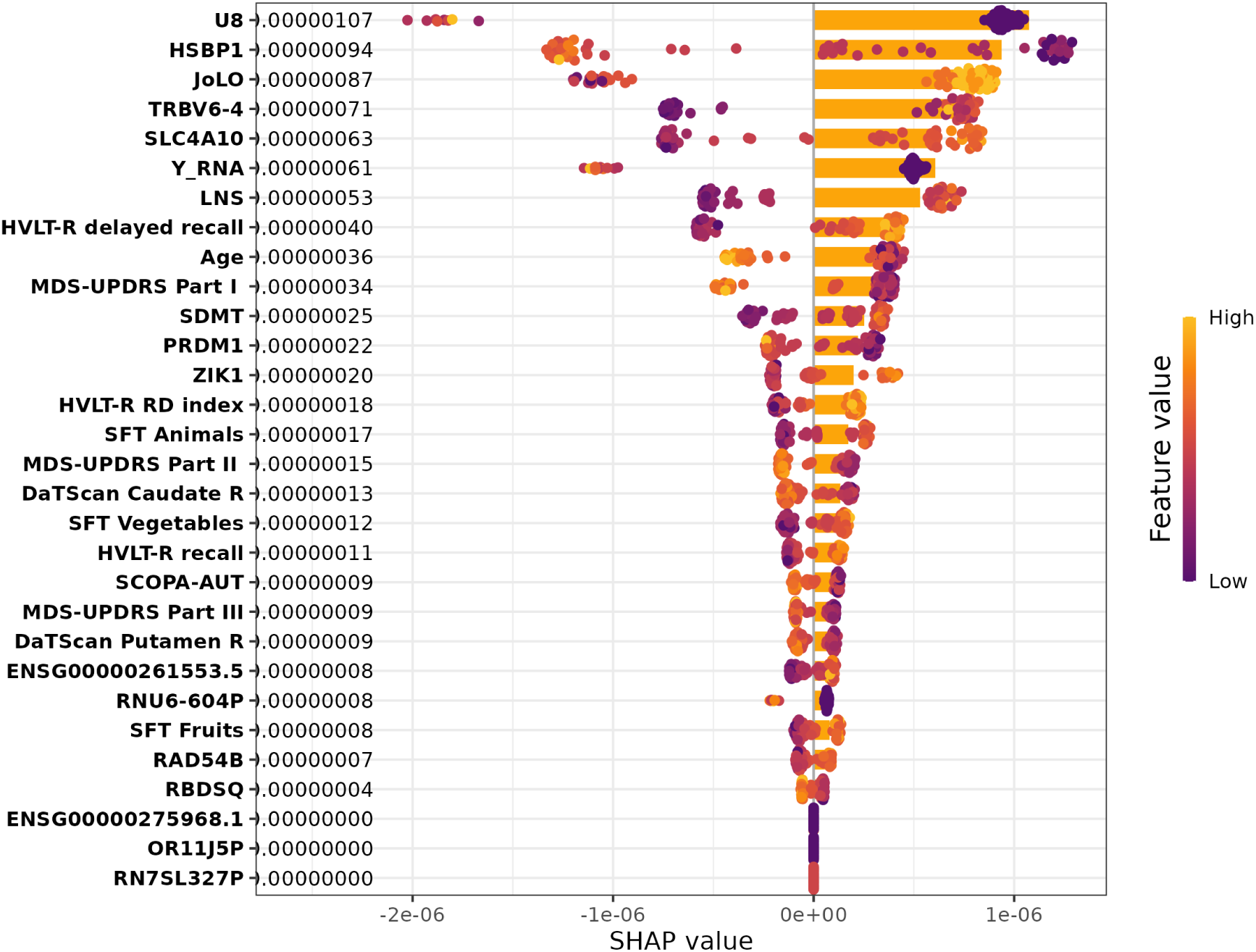
SHAP summary plot representing the contribution of each variable to the prediction of the model.

## 3 Discussion

The identification of progression subtypes is of extreme importance in order to attempt settling the heterogeneity of PD. Recent research has shown that people with PD can exhibit a variety of progression patterns from diagnosis onwards [5, 8, 19, 20, 22, 24]. The identification of disease-modifying treatments can be fostered by finer-grained diagnoses and biomarkers identification, pursuing a precision medicine approach. Targeting specific biological processes is currently unfeasible due to the lack of validated nonclinical biomarkers of PD progression [25], thus the importance of describing the biological profiles of progression subtypes is a paramount objective.

In this study we investigated the transcriptomic profile of three disease progression subtypes, which were identified in [20] with an Artificial Intelligence algorithm that reliably takes into account time as a dimension. Briefly, S1 had mild motor and non-motor symptoms at baseline, with a moderate rate of motor impairment increase and relatively stable cognitive abilities; S2 had moderate motor and non-motor symptoms at baseline, with a slow progression rate; and S3 started with significant motor and non-motor symptoms, showing a rapid progression of impairment, and thus reporting the worse prognosis among the three [20].

The DEGs identified in this study are unique to these progression subtypes, as none of the genes that are commonly found as differentially expressed in PD studies are present in our results. As a specific example, common transcriptomic markers such as *SYN1*, *ANKRD22*, and *SLC14A1* [16] are absent from all our DEGs lists. This result is not surprising to us, as our experiment had two main differences with other PD RNA studies. First, although based on transcriptomics of PD subjects, we investigated progression subtypes as diagnostic classes, thus differences with a classical PD group were expected. Second, our differential expression analysis was performed as a time course experiment, in order to identify those genes that varied for expression values as a result of the disease over time. This profoundly differs with previous PD transcriptomics studies, which performed a cross-sectional analysis of gene expression, thus not taking time into account. As a further note, there is general poor consensus between previous studies on resulting DEGs from PD studies [15].

### 3.1 S1

All pathway analyses consistently highlighted the modulation of cellular energy metabolism, particularly pathways associated with oxidative phosphorylation, aerobic respiration, and cellular respiration. Additionally, pathways related to ATP synthesis, mitochondrial dysfunction, and nucleotide metabolism were commonly enriched across the ORA and GSEA over GO, KEGG and WikiPathways databases. The modulation of energy metabolism is well known in PD, and it has already been found from transcriptomics analyses both in blood and brain sample tissues [26, 27, 28]. Cellular response to stress pathways, including oxidant detoxification and response to reactive oxygen species, were also consistently identified. Furthermore, the results consistently pointed towards the involvement of neurological diseases and neurodegeneration, with pathways associated with Parkinson’s disease, Alzheimer’s disease, Huntington disease, prion disease, and amyotrophic lateral sclerosis consistently enriched.

Despite these similarities, there were also differences observed across the pathway analyses of S1 data. In fact, there were different specific pathways within the broader common themes. For instance, one analysis emphasized the significance of pathways related to ribosomal proteins in protein synthesis, while another highlighted the importance of neuronal signaling pathways and immune system dysregulation. Disease-related pathways such as nonalcoholic fatty liver disease and hepatitis B infection were specifically enriched in one analysis. The involvement of immune system response and processes related to oxidative stress are known in PD transcriptomics [15, 17], and the observation of disease pathways enrichment is related to their modulation.

The biological profile of S1 shares similarities with that of PD patients with *LRRK2* mutation, which is involved in multiple biological functions, including mitochondrial activity and oxidative pathways [29]. It is interesting to note that none of the patients included in this study had a mutation in one of the risk loci known for PD, as this study was solely focused on idiopathic PD. Nonetheless, it has already been observed that the patients with idiopathic PD or *LRRK2* genetic PD show mostly overlapping phenotypes, and they are clinically difficult to distinguish [30].

Cellular signaling pathways were also found enriched in the GSEA, confirming that signaling mechanisms, often found among transcriptomics alterations from PD *post mortem* brain tissues [31], can also emerge from the analysis of peripheral tissues, such as blood [32, 33].

### 3.2 S2

Pathway analyses consistently identified modulation of gene expression regulation and metabolic processes. Specifically, pathways associated with RNA metabolism and processing emerged among the most significant terms across all analyses. The implication of RNA metabolic processes has been considered in the pathogenesis and disease course of PD, advancing that these may be related to energy conservation, aggregated proteins modulation, and response to cellular stress [34].

One notable difference lies in the number of pathways identified in each analysis, as some analyses revealed a limited number of pathways. As an example, there were only two significant pathways in the ORA on WikiPathways: Endoderm differentiation (WP2853) and Mesodermal commitment pathway (WP2857). These were enriched by a single gene, *NCAPG2*. This gene encodes for a regulatory subunit of the condensin II complex which, along with the condensin I complex, plays a role in chromosome assembly and segregation during mitosis [35]. Alterations of this gene have been associated with cancer and neurodevelopmental defects [36, 37], and although its presence has already been observed in PD blood transcriptomics [38, 39], its role in the disease is still unclear.

Cell-cell communication was found modulated in the GSEA results on all databases. Relatedly, various pathways related to stimulus response emerged as modulated, indicating their involvement in this phenotype.

In GSEA results on the KEGG database, S2 exhibited pathways associated with addiction processes, sharing this characteristic with S1. Pathways related to morphine addiction also emerged in a recent evaluation of PD proteome from dopaminergic neurons in the substantia nigra (SN), suggesting an involvement of potentially compromised GABA-related pathways [40].

### 3.3 S3

This subtype had significantly fewer shared terms with the other two, which in turn showed a much higher level of similarity. Pathways resulting from the ORA on DEGs indicate the involvement of response to oxidative stress and detoxification processes, aligning with findings in S1. Additionally, ORA on the KEGG database highlighted pathways related to diseases such as African trypanosomiasis and Malaria, implying a possible modulation of detoxification processes within this phenotype. Overall, these resulting pathways indicate a modulation of the processes associated with cellular adaptation and defense against oxidative stress and toxic substances. Cellular signaling was also found modulated in many of the results sets, and this is a shared alteration for all three subtypes. Accordingly, S3 results included sensory perception and signal transduction as prominent themes, with pathways related to the detection of chemical stimuli and smell perception. Also, the enrichment of pathways related to olfactory transduction, neuroactive ligand-receptor interaction, and protein export was observed. Furthermore, pathways associated with gene expression regulation and cellular response to misfolded proteins were significant, as also found in the other subtypes.

Metabolic pathways such as Vitamin B12 metabolism, Folate metabolism, and Selenium micronutrient network, were also found altered in this subtype. Recent studies have shown that B12 deficiency is common in patients with neuropathies, and PD has B12 levels decline over the course of the disease [41].

### 3.4 Results comparison between subtypes

The results of our transcriptomics analysis revealed a number of similarities between the three PD subtypes (S1, S2, and S3). All three subtypes showed a significant modulation of pathways related to the regulation of gene expression, metabolism, and cell signaling. Pathways associated with nervous system dysregulation were consistently found in all three subtypes. Although expected when analyzing brain cells, we believe that when resulting in blood it’s a confirmatory result of appropriate transcriptomics findings, and this is also in line with previous works on peripheral tissues [15, 42]. We may consider this as a general alteration due to the disease state, as these were also found in other PD transcriptomics experiments [16], and not distinctive of any of the subtypes.

S1 and S2 had a few shared themes, including addiction pathways, structure development, immune response alterations and disease processes. In fact, among the distinctive characteristics for S1 we find neurological and neurodegenerative disease pathways. Moreover, S1 was unique in its alteration of energy production and mitochondrial functions. Interestingly, all of the shared pathways between S1 and S2 had opposite enrichment patterns in the GSEA (Figure 6). This demonstrates that S1 and S2 are distinct progression forms of the same disease. Despite sharing a few transcriptomic characteristics, these appear to be modulated in opposing ways, and thus may be at the foundation of their different progression courses. S2 was unique in its alteration of pathways related to developmental processes and neurogenesis. Moreover, this subtype showed an alteration in olfactory transduction, as also observed in S3. S3 was unique in its increased expression of genes involved in detoxification processes, and pathways related to cellular stress response were altered in both S1 and S3. Interestingly, this was the only subtype characterized by enrichment of response to misfolded proteins.

### 3.5 Subtype prediction at baseline

The machine learning classifier provided a reliable tool to predict disease progression subtypes using baseline data. This tool could easily be implemented into a user-friendly software, to finally build a reliable Computer-Aided Diagnosis (CAD) tool to identify subjects with the most severe prognosis. As resulting from the variable importance analysis, the contribution of gene expression was marginal for the prediction of S3, not allowing for substantial discrimination between disease subtypes in neither of the steps of the hierarchical ML approach. Clinical variables instead demonstrated high importance to identify S3 subjects, with perceived disability (MDS-UPDRS Part II) being the most important predictor for a more severe prognosis. In fact, S3 subjects were characterized by a faster progression and worse symptomatology, sharing some similarities with the classical Posture Instability / Gait Difficulty (PIGD) subtype. Interestingly, most of the S3 subjects were PIGD patients, and those that were Tremor Dominant (TD) instead were likely to shift to PIGD over 6 years [20]. Although expression values resulted as the most important factors to discriminate between S1 and S2, the model at the second level of the hierarchy had a poor test performance. This made it unreliable and, as a consequence, the evaluation of its behavior is meaningless. Considering that this hierarchical classification model has 0.877 AUROC to detect the most severe subtype, this would give useful indication for prognosis. As such, this ML model may foster precision medicine for PD, providing support for a finer-grained diagnosis by applying the results of subtyping research. As all PD subjects included in this study were newly diagnosed, and the classifier was trained and tested on baseline data, it could be applied in clinical practice when evaluating a new PD patient. Additionally, we would like to highlight that the model was trained on baseline data to predict a class defined by disease progression, which involves the passage of time. Notably, it has a greater ability to predict a subject’s future compared to traditional PIGD/TD subtyping. This prediction holds particular relevance for individuals whose phenotype aligns with the S3 subtype, where this classification is more prone to change over time.

In the replication study of the PD progression subtype identification, it has been found that the most severe subtype (S3) had distinctive clinical features when compared to the two less severe subtypes (S1 and S2). Moreover, it was observed that there was limited signal in baseline variables to discriminate between the less severe subtypes [22]. These observations are in line with our results, as the performance of our classifier is poor in discriminating between S1 and S2 (0.576 AUROC). Additionally, our analysis revealed that not even transcriptomics assessment was useful to discriminate between S1 and S2 at baseline.

Providing a tool for progression subtype prediction at baseline is pivotal to improve the application of subtyping research results into PD clinical practice. Not only this study provides a biological characterization of progression subtypes, but it also demonstrates that a hierarchical ML approach is suitable to detect the most severe subtype, with a potentially relevant impact on prognosis.

### 3.6 Strengths and limitations

This study provides a characterization of the transcriptomics profile for three PD subtypes identified in a data-driven manner, namely using AI to analyze the disease progression. A datadriven approach to disease subtyping is free from the biases due to the experimenter and is more precise, as no a priori choices based on medical expertise are made. PPMI has one of the largest PD cohorts to date, offering a consistently large group to identify disease subtypes with AI methods. As the identification of disease progression subtypes was performed using an LSTM [20], the present study is hypothesis-free and aims to characterize the most reliable PD progression subtypes available in the literature.

The vastness of the results tables from the pathway analyses hindered results manageability. As a group of researchers, we did our best to read the results table and report noteworthy results, yet it is to be disclosed that a complete and accurate report was unfeasible. As a comment to this, we would like to speculate that future technological development may help with the interpretation of High Throughput Sequencing data analysis results: Large Language Models (LLM), such as ChatGPT [43], are showing increasingly better ability to handle textual data, and may one day be well-suited to summarize and expose these kinds of results. Potential future analysis of our results by means of such methods is encouraged, and full results tables can be found in Supplementary Tables 1-2.

## 4 Methods

### 4.1 Workflow overview

Data from the PPMI database were used for both of the objectives of this study: (1) to identify the transcriptomics characteristics of the disease progression subtypes, and (2) to train the ML model aimed at evaluating the usefulness of gene expression data in predicting disease subtypes at baseline. First, data were gathered and the cohort of study was defined, as described in section 4.2. RNA-Seq data were preprocessed (section 4.3) and then a differential expression analysis was performed as described in section 4.4 The resulting DEGs were further analyzed through pathway analyses, as described in section 4.5 Following cohort definition, the ML classifier was trained to predict the cluster at baseline, as described in section 4.6, then its behaviour was investigated using XAI methods (section 4.7). R code used to perform data analysis can be found on GitHub (https://github.com/217c/parkinson_subtypes_rnaseq, accessed on 25 September 2023).

### 4.2 Data

Data used in this study were obtained from the Parkinson Progression Marker Initiative (PPMI) [44]. PPMI is one of the most important ongoing studies of PD progression markers, collecting data from multiple international sources and focusing on a diverse range of potential markers for tracking the progression of PD, including demographics, clinical variables, imaging data, cerebrospinal fluid, blood, DNA and, importantly to this study, RNA measures. The data were downloaded from the LONI Image and Data Archive (IDA) in April 2022. The cohort of study was defined using the PPMI Consensus Committee Analytic Dataset (RD: 2021-10-28). PPMI inclusion criteria for PD subjects include: age ≥ 30, Parkinson’s disease diagnosis within the last 2 years, baseline Hoehn and Yahr Stage I–II, and no anticipated need for symptomatic treatment within 6 months of baseline [44]. Healthy controls (HC) inclusion criteria will include individuals without clinical signs suggestive of parkinsonism, no evidence of cognitive impairment, and no first-degree relative diagnosed with PD. To be included in this study cohort, subjects must have had a diagnosis of sporadic PD and available RNA-Seq data for multiple timepoints, as found in the LongRNA Transcriptome Sequencing of PPMI Samples (B38) study (RD: 2021-04-02). The PPMI RNA Sequencing Project has generated overview transcriptomics data from raw sequencing reads of PPMI whole blood samples. The data were pre-analyzed and quality-controlled from the PPMI group [45].

The definition of the sample for this study follows that described in [20]. Subjects that underwent disease progression subtyping were included, along with all available HC subjects. In brief, S1 starts with mild motor and non-motor symptoms, and motor impairment increases with a moderate rate over time; S2 has moderate motor and non-motor symptoms at baseline, with a slow progression rate; S3 has significant motor and non-motor deficits at baseline, and its impairment progresses rapidly over time, thus accounting for a worse prognosis. The IDs of the subjects assigned to disease progression subtypes were retrieved from [20]. To summarize, data analysis was performed on those subjects that had RNA-Seq data available and that were clustered into one PD subtype. This study cohort included a total number of 2085 RNA-Seq records for 4 years of longitudinal measures (starting from baseline, with constant time interval measures at 12 months) from 600 subjects (PD = 407, HC = 193) (S1 = 199; S2 = 52; S3 = 156).

### 4.3 RNA-Seq data preparation

To assess outliers, a Principal Component Analysis (PCA) was computed on variance stabilized and transformed (namely, vst from DESeq2) expression data of the top 20000 genes, and data points lying beyond the edges of the Highest Density Interval of the first principal component were deemed as outliers. The threshold was set to 0.99, thus considering as outliers all observations outside the 99% CI [46]. A sex incompatibility check was performed to assess contamination due to abnormal transcription using t-SNE and DBSCAN on gene expression data from the following sex chromosome genes: *USP9Y, XIST, RPS4Y1, RPS4Y2, KDM5D, DDX3Y*. Subjects whose samples had inconsistent clustering between sex in metadata and sex from expression data were removed from the analysis (Supplementary Figure 1).

### 4.4 Differential Expression Analysis

Differentially expressed genes (DEGs) were identified using `DESeq2` R library v1.38.3 to perform a Likelihood ratio test (LRT). This experiment was designed as a time course analysis, thus the full model including group, time, and their interaction, was compared to a reduced model without the interaction. This analysis allowed us to identify those genes that at one or more time points after time 0 showed a group-specific effect, thus excluding genes that moved up or down in time in the same way in both groups. Each PD cluster was compared to the HC group performing a separated LRT. For each comparison, DESeq2 automatically estimated size factors based on the median ratio method, estimated dispersions, and performed the LRT for negative binomial GLMs [47]. Correction for multiple testing was performed using the False Discovery Rate (FDR) method, applying DESeq2’s default threshold for adjusted p-value < 0.1. Gene names and descriptions were retrieved using `g:Profiler` R package [48].

### 4.5 Pathway analysis

To further investigate the differences in gene expression we performed a pathway analysis using `clusterProfiler` R library v4.6.2 [49]. An Over-Representation Analysis (ORA) was performed on DEGs for all three comparisons on GO Biological Process (BP) domain, KEGG, and WikiPath-ways databases. Not to limit our pathway analysis to DEGs sets, we chose to investigate pathway modulation due to eventually small but coordinated changes in the expression levels of all genes, thus performing a Gene Set Enrichment Analysis (GSEA) for all three comparisons on GO BP, KEGG, and WikiPathways databases. To improve interpretability, GSEA results on GO were reduced to semantically similar terms using `rrvgo` R library v1.10.0 [50].

### 4.6 Machine Learning model for subtype prediction at baseline

Data collected at the time of diagnosis (baseline) was used to predict the cluster, using a hierarchical machine learning approach. In this approach, we train multiple classifiers in a hierarchical structure, where each classifier is responsible for a specific task. This approach is useful here because the classification task can be broken down into simpler sub-tasks. As cluster 3 showed to be the most severe, the first step was to predict if the newly diagnosed PD subject belonged to S3. If not, the second step aimed to predict whether the subject was from S1 or S2 (Figure 13).

**Figure 13:**
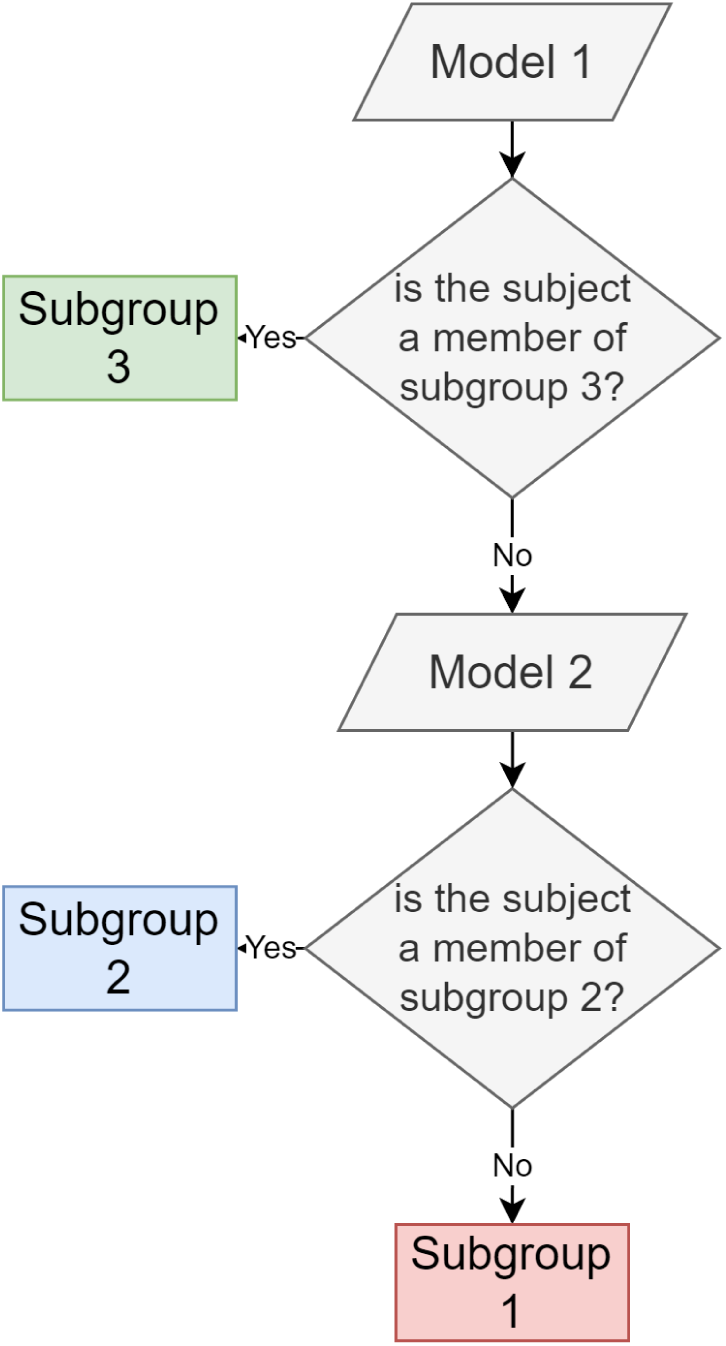
Schematic representation of the flow of the Hierarchical ML approach.

Specifically, two XGBoost models were used in this pipeline [51]. Firstly, a subject is evaluated by the first model in the hierarchy, which aims to identify subjects from S3. If the subject is found to be from S3, the pipeline ends. If the subject is found not to be from S3, then the subject is evaluated by the second model, which aims to discriminate between S2 and S1 subjects. The machine learning pipeline was developed using `tidymodels` R library v1.0.0. Train test split was performed at subject level, including 75% of the sample in the train set (Table 1). Data from multiple modalities were used, including demographics, motor, non-motor, biospecimen, imaging, and gene expression values (Table 2). Missing data were imputed with the mean value of the train set and rounded to integer value, thus respecting the original format of variables. All variables were transformed by applying a Box-Cox transformation [52] and feature selection was performed by univariate filtering with ANOVA on all three groups. Variables reporting an FDR-corrected p-value < 0.05 were selected for training. Variables with an absolute Pearson’s correlation value greater than 0.8 with other variables were removed. Synthetic minority oversampling technique (SMOTE) was used to address class imbalance before training [53]. The XGBoost models were trained using 10 Cross-Validation resamples to find the best combination of hyperparameters using a grid latin hypercube of values [54]. The best models resulting from cross-validation were tested on the test set and evaluation metrics were computed.

**Table 1:** Number of observations in train and test splits.

| Split | Subtype | n |
| --- | --- | --- |
| train | S1 | 141 |
| train | S2 | 33 |
| train | S3 | 108 |
| test | S1 | 47 |
| test | S2 | 12 |
| test | S3 | 37 |

**Table 2:**
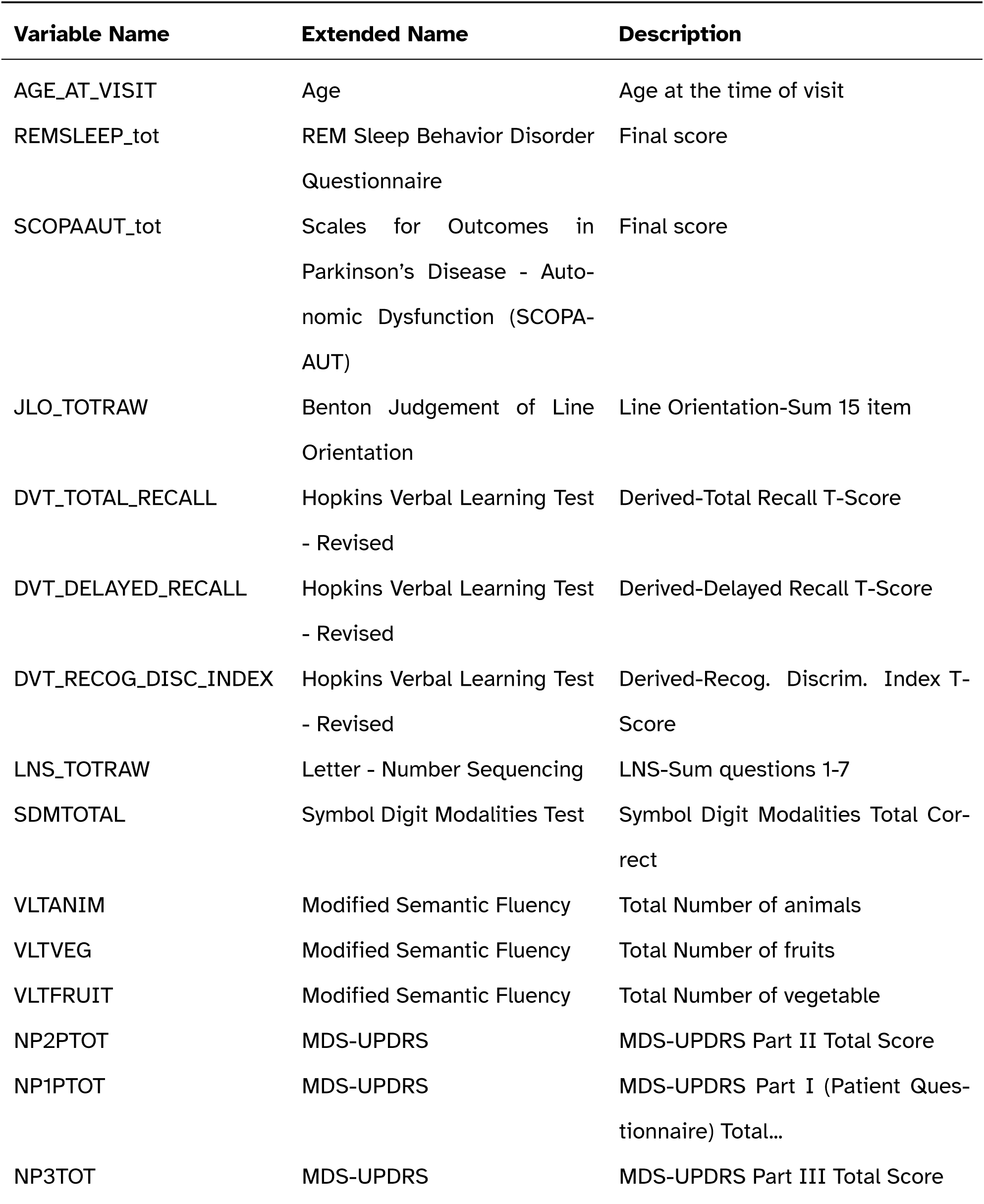
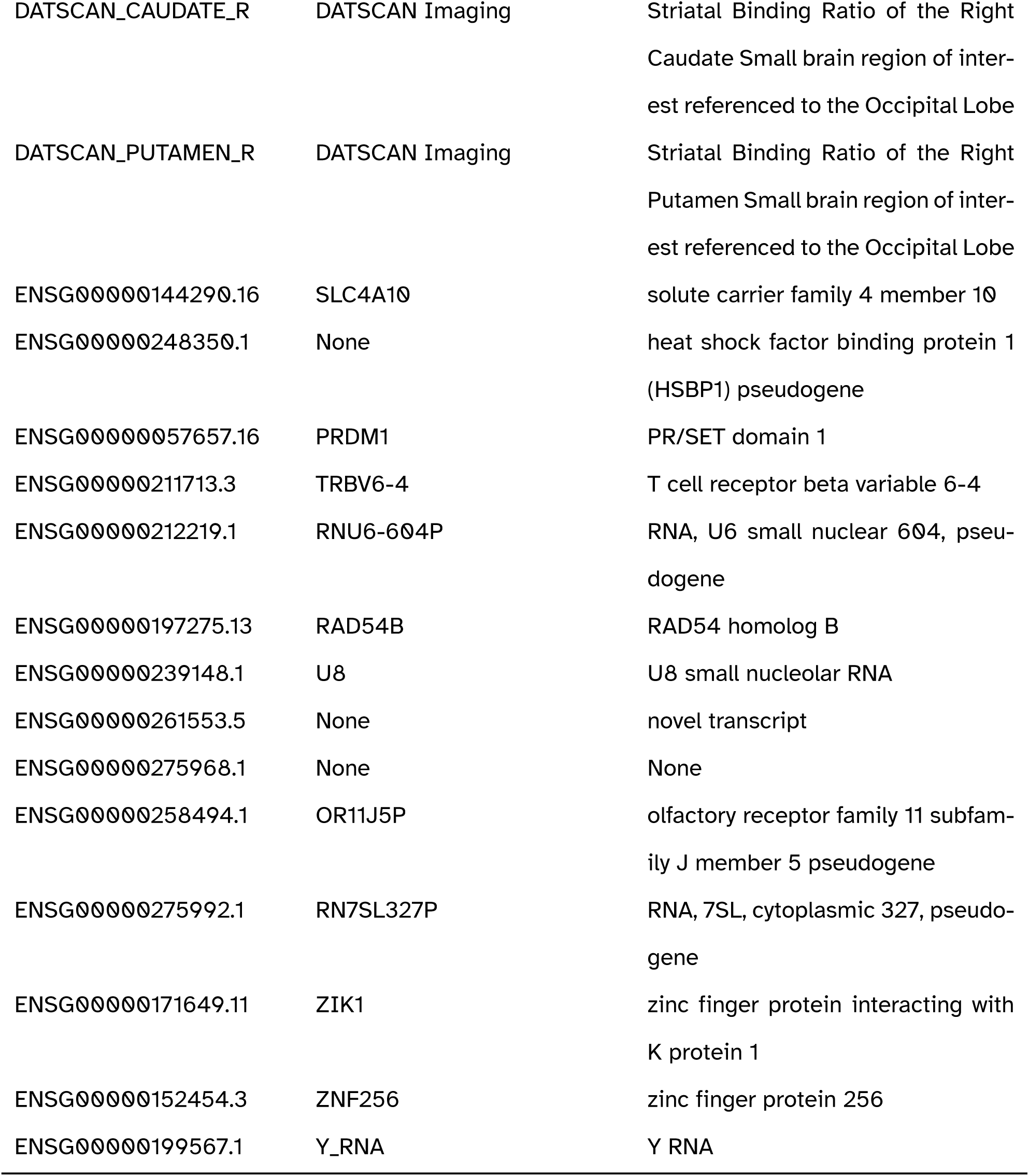
Full list of variables used for machine learning.

### 4.7 Variable importance and XAI

The importance of variables in contributing to the Machine Learning prediction of subtype at baseline was investigated using SHAP (SHapley Additive exPlanations) values [55]. As an XAI method [56], SHAP values highlight the contribution of each feature to the final prediction, thus providing a measure to rank features importance. To calculate SHAP values and produce informative plots, `shapviz` R library functions [57] were applied to the XGBoost models.

## Data Availability

Data used in the preparation of this article were obtained from the Parkinson's Progression Markers Initiative (PPMI) database (www.ppmi-info.org/access-data-specimens/download-data), RRID:SCR_006431.
The PPMI IDs of the subjects in the disease subtypes were obtained from the GitHub repository related to the following article: Zhang, X., Chou, J., Liang, J. et al. Data-Driven Subtyping of Parkinson's Disease Using Longitudinal Clinical Records: A Cohort Study. Sci Rep 9, 797 (2019). https://doi.org/10.1038/s41598-018-37545-z

## Data availability statement

Data used in the preparation of this article were obtained from the Parkinson’s Progression Markers Initiative (PPMI) database (www.ppmi-info.org/access-data-specimens/download-data), RRID:SCR_006431. The PPMI IDs of the subjects in the disease subtypes were obtained from the GitHub repository related to [20].

## Acknowledgements

PPMI – a public-private partnership – is funded by the Michael J. Fox Foundation for Parkinson’s Research and funding partners, including 4D Pharma, Abbvie, AcureX, Allergan, Amathus Therapeutics, Aligning Science Across Parkinson’s, AskBio, Avid Radiopharmaceuticals, BIAL, Biogen, Biohaven, BioLegend, BlueRock Therapeutics, Bristol-Myers Squibb, Calico Labs, Celgene, Cerevel Therapeutics, Coave Therapeutics, DaCapo Brainscience, Denali, Edmond J. Safra Foundation, Eli Lilly, Gain Therapeutics, GE HealthCare, Genentech, GSK, Golub Capital, Handl Therapeutics, Insitro, Janssen Neuroscience, Lundbeck, Merck, Meso Scale Discovery, Mission Therapeutics, Neurocrine Biosciences, Pfizer, Piramal, Prevail Therapeutics, Roche, Sanofi, Servier, Sun Pharma Advanced Research Company, Takeda, Teva, UCB, Vanqua Bio, Verily, Voyager Therapeutics, the Weston Family Foundation and Yumanity Therapeutics.

For up-to-date information on the study, visit www.ppmi-info.org.

