## Supplementary Figure 1 for "Transcriptomics profiling of Parkinson’s disease progression subtypes reveals distinctive patterns of gene expression"

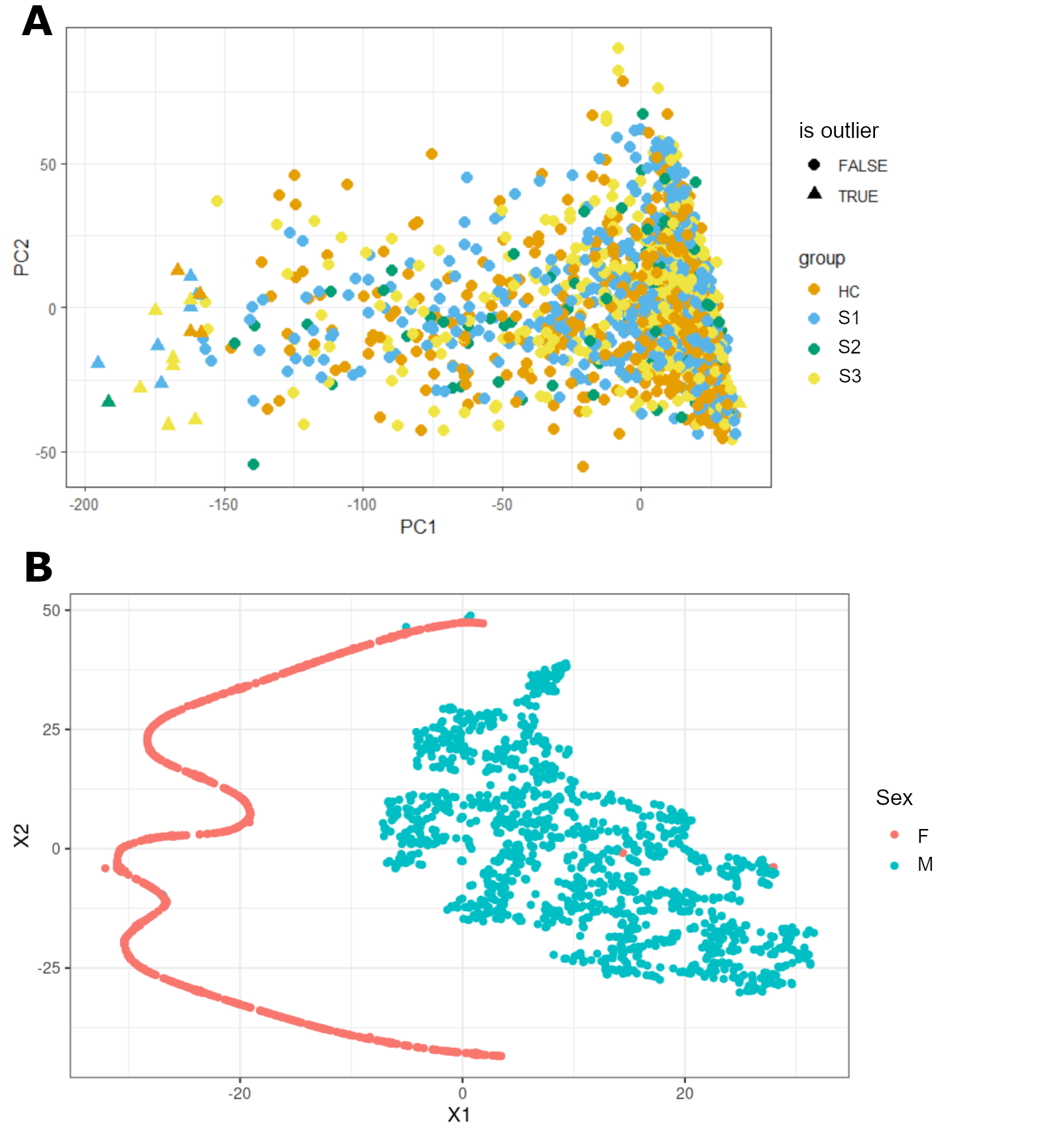

Supplementary Figure 1 - A) PCA components 1 and 2 of the top 20000 genes. Outlier samples were identified at the edges of the distribution, and are represented as triangles.

B) TSNE plot of gene expression values for six sex chromosome genes. Samples are well separated upon sex factor, and samples showing inconsistent clustering were removed from the analysis.
